# Effectiveness of BNT162b2 and mRNA-1273 COVID-19 vaccines against symptomatic SARS-CoV-2 infection and severe COVID-19 outcomes in Ontario, Canada: a test-negative design study

**DOI:** 10.1101/2021.05.24.21257744

**Authors:** Hannah Chung, Siyi He, Sharifa Nasreen, Maria E. Sundaram, Sarah A. Buchan, Sarah E. Wilson, Branson Chen, Andrew Calzavara, Deshayne B. Fell, Peter C. Austin, Kumanan Wilson, Kevin L. Schwartz, Kevin A. Brown, Jonathan B. Gubbay, Nicole E. Basta, Salaheddin M. Mahmud, Christiaan H. Righolt, Lawrence W. Svenson, Shannon E. MacDonald, Naveed Z. Janjua, Mina Tadrous, Jeffrey C. Kwong, on behalf of the Canadian Immunization Research Network (CIRN) Provincial Collaborative Network (PCN) Investigators

## Abstract

**Objectives:** To estimate the effectiveness of mRNA COVID-19 vaccines against symptomatic infection and severe outcomes.

**Design:** We applied a test-negative design study to linked laboratory, vaccination, and health administrative databases, and used multivariable logistic regression adjusting for demographic and clinical characteristics associated with SARS-CoV-2 and vaccine receipt to estimate vaccine effectiveness (VE) against symptomatic infection and severe outcomes.

**Setting:** Ontario, Canada between 14 December 2020 and 19 April 2021.

**Participants:** Community-dwelling adults aged ≥16 years who had COVID-19 symptoms and were tested for SARS-CoV-2.

**Interventions:** Pfizer-BioNTech’s BNT162b2 or Moderna’s mRNA-1273 vaccine.

**Main outcome measures:** Laboratory-confirmed SARS-CoV-2 by RT-PCR; hospitalization/death associated with SARS-CoV-2 infection.

**Results:** Among 324,033 symptomatic individuals, 53,270 (16.4%) were positive for SARS-CoV-2 and 21,272 (6.6%) received ≥1 vaccine dose. Among test-positive cases, 2,479 (4.7%) had a severe outcome. VE against symptomatic infection ≥14 days after receiving only 1 dose was 60% (95%CI, 57 to 64%), increasing from 48% (95%CI, 41 to 54%) at 14–20 days after the first dose to 71% (95%CI, 63 to 78%) at 35–41 days. VE ≥7 days after 2 doses was 91% (95%CI, 89 to 93%). Against severe outcomes, VE ≥14 days after 1 dose was 70% (95%CI, 60 to 77%), increasing from 62% (95%CI, 44 to 75%) at 14–20 days to 91% (95%CI, 73 to 97%) at ≥35 days, whereas VE ≥7 days after 2 doses was 98% (95%CI, 88 to 100%). For adults aged ≥70 years, VE estimates were lower for intervals shortly after receiving 1 dose, but were comparable to younger adults for all intervals after 28 days. After 2 doses, we observed high VE against E484K-positive variants.

**Conclusions:** Two doses of mRNA COVID-19 vaccines are highly effective against symptomatic infection and severe outcomes. Single-dose effectiveness is lower, particularly for older adults shortly after the first dose.

## INTRODUCTION

Understanding how clinical trial efficacy estimates of COVID-19 vaccines translate into real-world effectiveness estimates is crucial, given differences in populations, dosing intervals, and emerging variants.[1] Due to COVID-19 vaccine supply constraints, Canada’s National Advisory Committee on Immunization (NACI) recommended extending the interval between doses to a maximum of 16 weeks.[2] With vaccine supply constraints globally, determining the effectiveness of these vaccines following a single dose vs. two doses is important for informing policy for many countries.[1]

We applied the test-negative design to linked, population-based health databases in Ontario, Canada (population 15 million) to evaluate vaccine effectiveness (VE) against symptomatic SARS-CoV-2 infection and severe outcomes (i.e., hospitalization or death associated with SARS-CoV-2 infection) for two mRNA vaccines (Pfizer-BioNTech’s BNT162b2 and Moderna’s mRNA-1273).

## METHODS

### Study population, setting, and design

We conducted a test-negative design study among community-dwelling Ontarians who had symptoms consistent with COVID-19. The test-negative design is comparable to a nested case-control design, with symptomatic individuals who are tested for the presence of a pathogen of interest serving as the nesting cohort.[1,3,4] All Ontarians aged ≥16 years, eligible for provincial health insurance, not living in long-term care, and who were tested for SARS-CoV-2 between 14 December 2020 and 19 April 2021 were eligible for inclusion. We excluded individuals who tested positive for SARS-CoV-2 prior to 14 December 2020 and recipients of the ChAdOx1 vaccine. We restricted the analysis to individuals who had at least one relevant COVID-19 symptom (based on self-report or observation [e.g., measured temperature]) at the time of testing, which was collected on the SARS-CoV-2 test requisition (see **Supplementary Methods**). Although these individuals and other members of the general public contributed in important ways to this research, it was not feasible to involve study subjects or the public in the design, conduct, reporting, or dissemination plans of our research.

### Data sources and definitions

We linked data from provincial SARS-CoV-2 laboratory testing, COVID-19 vaccination, and health administrative datasets using unique encoded identifiers and analyzed them at ICES (formerly the Institute for Clinical Evaluative Sciences).

#### Outcomes

Our first primary outcome was symptomatic SARS-CoV-2 infection, ascertained by real-time reverse transcription polymerase chain reaction (RT-PCR) tests on respiratory specimens, including nasopharyngeal (most common), nasal, throat, saliva, and turbinate.[5] Using data from the Ontario Laboratories Information System (OLIS), which captured 91.8% (n=258,207) of all provincially reported cases of laboratory-confirmed COVID-19 (n=281,261) during the study period, test-positive individuals were treated as cases and test-negative individuals were treated as controls. Since symptom onset dates were inconsistently reported in OLIS, we used the specimen collection date as the index date. For cases with multiple positive tests, we used the date of their first positive test. For controls with multiple negative tests, we used the date of a randomly selected negative test as the index date.

We obtained information on variants and mutations from the Public Health Case and Contact Management system (CCM), which contains information on the clinical course of cases and the results of screening tests for N501Y and E484K mutations and whole genome sequencing results that identify specific variant of concern (VOC) lineages (Alpha [B.1.1.7], Beta [B.1.351], Gamma [P.1]). All RT-PCR-positive specimens with cycle threshold values ≤35 were tested for the N501Y mutation (starting 3 February 2021) and the E484K mutation (starting 22 March 2021).[6] We considered samples with positive N501Y and negative E484K mutations as Alpha, and samples with positive N501Y and E484K mutations as Beta or Gamma. We combined the latter two lineages for our analysis because there were very small numbers of cases identified using whole genome sequencing.

Our second primary outcome was severe disease associated with symptomatic SARS-CoV-2 infection, defined as either hospitalization or death with a recent positive test, using the earliest of the specimen collection date or the hospitalization or death date as the index date. We identified these outcomes using CCM (for both hospitalizations and deaths), the Canadian Institute for Health Information’s Discharge Abstract Database (DAD; for hospitalizations), and the Ontario Registered Persons Database (RPDB; for deaths). For hospitalizations identified using DAD, a positive test must have occurred within 14 days prior to or 3 days after admission. For deaths identified using RPDB, a positive test must have occurred within 30 days prior to death or within 7 days post-mortem. We used the same control group as the first primary outcome analysis (i.e., symptomatic individuals who tested negative for SARS-CoV-2).

#### COVID-19 vaccination

BNT162b2 became available in Ontario on 14 December 2020, and mRNA-1273 on 28 December 2020.[7] The initial vaccination phase prioritized high-risk populations such as older adults living in congregate settings, healthcare workers (including non-patient-facing staff working in healthcare institutions), adults living in Indigenous communities, and adults aged ≥80 years.[7] Ontario had initially followed the manufacturers’ recommended dosing schedules (i.e., a 21-day interval for BNT162b2 and a 28-day interval for mRNA-1273), but due to disruptions in vaccine supply in late January 2021, extended the interval to 35-42 days for everyone except older adults living in congregate settings and Indigenous individuals. In early March, Ontario adopted NACI’s recommendation to delay administration of the second dose by up to 16 weeks for most individuals.[8,9] Eligibility expanded over time, taking into account both age (i.e., graduated expansion by decreasing age) and other high-risk populations, such as individuals with certain health conditions and their caregivers, certain essential frontline workers, and those aged ≥18 years living or working in high-COVID-19-incidence communities (i.e., those disproportionately affected by COVID-19 and where transmission was still high). Adherence to these eligibility criteria varied across regions. As of 19 April 2021, 28% of Ontario adults had received at least one dose of a COVID-19 vaccine.[10] Comprehensive documentation of all COVID-19 vaccination events in Ontario, including product, date of administration, and dose number, is recorded in real-time into COVaxON, a centralized COVID-19 vaccine information system. We used the COVaxON file containing events up to 25 April 2021 for these analyses, which likely had records of all vaccinations delivered by 19 April 2021.

#### Covariates

We obtained age, sex, and postal code of residence as of 14 December 2020 from RPDB. We obtained the number of RT-PCR tests for each subject during the 3 months prior to 14 December 2020 from OLIS to use as a proxy for highly tested individuals at increased risk of exposure to SARS-CoV-2 infection (e.g., healthcare workers and caregivers of long-term care residents, who must also undergo serial SARS-CoV-2 testing). We grouped testing dates into 2-week periods to capture temporal changes in viral activity and regional vaccine roll-out. We determined the presence of comorbidities that increase the risk of severe COVID-19,[11] identified from various databases using validated algorithms and commonly accepted diagnostic codes, which have been described elsewhere.[12] We ascertained receipt of influenza vaccination during the 2019/2020 and/or 2020/2021 influenza season (a proxy for health behaviours) using physician and pharmacist billing claims in the Ontario Health Insurance Plan and Ontario Drug Benefit databases, respectively. We determined the public health unit of residence using the postal code and Statistics Canada Postal Code Conversion File Plus (version 7B) and grouped them into larger regions. We obtained information at the ecologic level of dissemination area (DA) on four important social determinants of health (median household income, proportion of the working population employed as non-health essential workers [i.e., those unable to work from home], average number of persons per dwelling, and proportion of the population who self-identify as a visible minority) from 2016 Census data.[13]^13^ DAs generally contain 400-700 individuals. Details related to these covariates are available in **eTable 1**.

### Statistical analysis

We conducted descriptive analyses and calculated standardized differences to compare characteristics between test-positive cases and test-negative controls, and between vaccinated and unvaccinated individuals.

We used multivariable logistic regression models to estimate the odds ratio (OR) comparing the odds of vaccination between test-positive cases and test-negative controls (where unvaccinated individuals served as the reference group). We estimated unadjusted and adjusted odds ratios accounting for all covariates listed above. These covariates were selected *a priori* based on their known associations with SARS-CoV-2 infection or severity and COVID-19 vaccine receipt,[2,11,14] and were assessed as potential confounders (**eTable 2**).[15] VE was calculated using the following formula: VE = (1-OR) x 100%. Individuals without information on exposures, outcomes, or covariates in ICES’ data holdings were assumed not to have the exposure, outcome, or covariate, and were categorized as such within the analyses.

For the primary analysis, we estimated overall VE (for both mRNA vaccines combined) for those who received only 1 dose by their index date and those who received 2 doses by their index date. We considered index dates within varying intervals after vaccination.

We also estimated VE ≥14 days after the first dose (among those who only received 1 dose) and ≥7 days after the second dose,[16] stratified by vaccine product (BNT162b2 or mRNA-1273), age group (16–39, 40–69, and ≥70 years), sex, presence of any comorbidity, epidemic wave (index dates 14 December 2020–7 February 2021, representing wave 2 in Ontario; 8 February 2021–21 March 2021, representing the period between wave 2 and wave 3; and 22 March 2021–19 April 2021, representing wave 3), and variant (earlier variant vs. Alpha vs. Beta or Gamma). We also estimated VE by varying intervals after vaccination stratified by age group.

We repeated these analyses for severe outcomes, with adjustments to the intervals after vaccination due to reduced sample sizes. For example, we evaluated VE for the entire period (i.e., ≥0 days) after receipt of the second dose.

Finally, as a sensitivity analysis, we determined VE against symptomatic infection and severe outcomes by varying intervals among only individuals who were vaccinated (treating individuals vaccinated 0-13 days before testing as the reference group) to assess whether systematic differences between vaccinated and unvaccinated individuals in the main analyses were adequately controlled for.[17]

All analyses were conducted using SAS Version 9.4 (SAS Institute Inc., Cary, NC). All tests were two-sided and used p*<*0.05 as the level of statistical significance.

## RESULTS

From 14 December 2020 – 19 April 2021, 2,171,449 unique individuals were tested for SARS-CoV-2. After excluding individuals who had SARS-CoV-2 infection prior to the study period and individuals who had received ChAdOx1 vaccine, 60.5% of those remaining did not have symptoms consistent with COVID-19 or had no symptom information recorded in OLIS, 24.4% were recorded as asymptomatic, and 15.1% had symptoms consistent with COVID-19 recorded at the time of testing (**Figure 1**). Grouped together, individuals with COVID-19-like symptoms and those deemed asymptomatic had similar characteristics as the remaining individuals, except for COVID-19 vaccine uptake, public health unit region, and number of previous SARS-CoV-2 tests (**eTable 3**).

**Figure 1:**
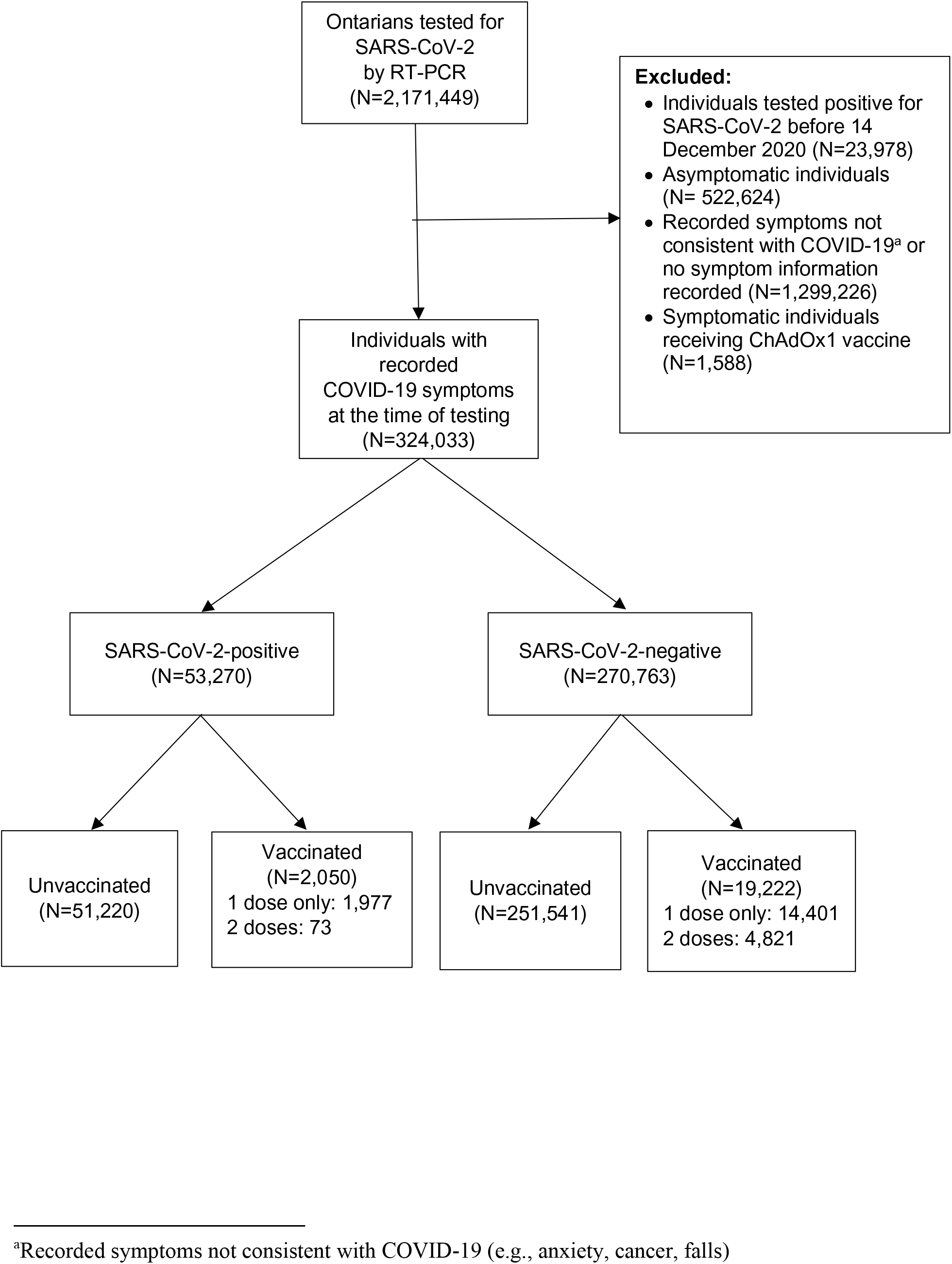
Community-dwelling adults included in the tested cohort between 14 December 2020 and 19 April 2021 in Ontario, Canada.

Of the 324,033 symptomatic tested individuals, 53,270 (16.4%) tested positive for SARS-CoV-2, 42,567 (79.9%) had variant testing information available, 21,272 (6.6%) had received ≥1 dose of mRNA vaccine, and 4,894 (1.5%) had received 2 doses (**Table 1**). Among test-positive cases, 2,479 (4.7%) had a severe outcome, of which 2,035 were hospitalized and 444 had died. Most hospitalized cases tested positive prior to or on admission date (1,728 [84.9%]) and nearly all tested positive before death. Test-positive cases were more likely to be male; more likely to reside in Peel Region or Toronto; more likely to have had zero SARS-CoV-2 tests during the 3 months prior to the vaccination program; less likely to have received an influenza vaccine; and more likely to reside in neighbourhoods with lower income, more persons per dwelling, and greater proportions of essential workers and visible minorities (**Table 1**). Vaccinated individuals were older; less likely to be male; more likely to have had multiple SARS-CoV-2 tests during the 3 months prior to the vaccination program; more likely to have a comorbidity; and more likely to have received an influenza vaccine. Compared to recipients of mRNA-1273 vaccine, recipients of BNT162b2 vaccine were younger, more likely to be female, and less likely to have a comorbidity (**eTable 4**). The distribution of these vaccine products also differed by Public Health Unit regions. Most individuals (77% for BNT162b2, 76% for mRNA-1273) had received only 1 dose by the index date. Distribution of vaccine product over the study period is presented in **eFigure 1.**

**Table 1.** Characteristics of symptomatic individuals tested for SARS-CoV-2 between 14 December 2020 and 19 April 2021 in Ontario, Canada.

| Characteristic | SARS-CoV-2-positive,<br>n (%)<sup>a</sup><br>(N=53,270) | SARS-CoV-2-negative,<br>n (%)<sup>a</sup><br>(N=270,763) | Standardized<br>difference<sup>b</sup> | Vaccinated with ≥1<br>dose of mRNA<br>COVID-19 vaccine,<br>n (%)<sup>a</sup><br>(N=21,272) | Unvaccinated,<br>n (%)<sup>a</sup><br>(N=302,761) | Standardized<br>difference<sup>b</sup> |
| --- | --- | --- | --- | --- | --- | --- |
| Received ≥1 dose of COVID-19 vaccine | 2,050 (3.8) | 19,222 (7.1) | - | - | - | - |
| Received 2 doses of COVID-19 vaccine | 73 (0.1) | 4,821 (1.8) | - | - | - | - |
| Tested positive for SARS-CoV-2 | - | - | - | 2,050 (9.6) | 51,220 (16.9) | - |
| Earlier variant | - | - | - | 579 (2.7) | 27,510 (9.1) | - |
| Alpha (B.1.1.7) | - | - | - | 807 (3.8) | 12,282 (4.1) | - |
| Beta (B.1.351) or Gamma (P.1)<br>(E484K+ variants) | - | - | - | 98 (0.5) | 1,291 (0.4) | - |
| Age (years), mean (standard deviation) | 42.4 (17.1) | 43.2 (17.8) | 0.04 | 51.8 (20.8) | 42.4 (17.3) | 0.49 |
| Age group (years) |  |  |  |  |  |  |
| 16–29 | 15,175 (28.5) | 72,238 (26.7) | 0.04 | 3,457 (16.3) | 83,956 (27.7) | 0.28 |
| 30–39 | 10,024 (18.8) | 59,326 (21.9) | 0.08 | 4,011 (18.9) | 65,339 (21.6) | 0.07 |
| 40–49 | 9,642 (18.1) | 46,225 (17.1) | 0.03 | 3,287 (15.5) | 52,580 (17.4) | 0.05 |
| 50–59 | 9,460 (17.8) | 40,874 (15.1) | 0.07 | 3,112 (14.6) | 47,222 (15.6) | 0.03 |
| 60–69 | 5,279 (9.9) | 27,342 (10.1) | 0.01 | 2,261 (10.6) | 30,360 (10.0) | 0.02 |
| 70–79 | 2,426 (4.6) | 14,888 (5.5) | 0.04 | 2,272 (10.7) | 15,042 (5.0) | 0.21 |
| ≥80 | 1,264 (2.4) | 9,870 (3.6) | 0.07 | 2,872 (13.5) | 8,262 (2.7) | 0.40 |
| Male sex | 25,993 (48.8) | 112,501 (41.5) | 0.15 | 6,013 (28.3) | 132,481 (43.8) | 0.33 |
| Public health unit region<sup>c</sup> |  |  |  |  |  |  |
| Central East | 2,624 (4.9) | 29,194 (10.8) | 0.22 | 1,969 (9.3) | 29,849 (9.9) | 0.02 |
| Central West | 8,322 (15.6) | 48,419 (17.9) | 0.06 | 3,853 (18.1) | 52,888 (17.5) | 0.02 |
| Durham | 1,433 (2.7) | 8,583 (3.2) | 0.03 | 522 (2.5) | 9,494 (3.1) | 0.04 |
| Eastern | 689 (1.3) | 15,147 (5.6) | 0.24 | 1,087 (5.1) | 14,749 (4.9) | 0.01 |
| North | 1,753 (3.3) | 31,321 (11.6) | 0.32 | 2,251 (10.6) | 30,823 (10.2) | 0.01 |
| Ottawa | 417 (0.8) | 3,144 (1.2) | 0.04 | 446 (2.1) | 3,115 (1.0) | 0.09 |
| Peel | 13,515 (25.4) | 32,981 (12.2) | 0.34 | 2,395 (11.3) | 44,101 (14.6) | 0.10 |
| South West | 7,562 (14.2) | 39,316 (14.5) | 0.01 | 3,885 (18.3) | 42,993 (14.2) | 0.11 |
| Toronto | 12,458 (23.4) | 45,540 (16.8) | 0.16 | 3,462 (16.3) | 54,536 (18.0) | 0.05 |
| York | 4,278 (8.0) | 15,995 (5.9) | 0.08 | 1,323 (6.2) | 18,950 (6.3) | 0.00 |
| Biweekly period of test |  |  |  |  |  |  |
| 14 Dec 2020 to 27 Dec 2020 | 4,139 (7.8) | 27,456 (10.1) | 0.08 | 13 (0.1) | 31,582 (10.4) | 0.48 |
| 28 Dec 2020 to 10 Jan 2021 | 6,870 (12.9) | 26,993 (10.0) | 0.09 | 335 (1.6) | 33,528 (11.1) | 0.40 |
| 11 Jan 2021 to 24 Jan 2021 | 4,864 (9.1) | 26,747 (9.9) | 0.03 | 1,068 (5.0) | 30,543 (10.1) | 0.19 |
| 25 Jan 2021 to 7 Feb 2021 | 3,539 (6.6) | 24,276 (9.0) | 0.09 | 1,204 (5.7) | 26,611 (8.8) | 0.12 |
| 8 Feb 2021 to 21 Feb 2021 | 3,595 (6.7) | 24,800 (9.2) | 0.09 | 1,031 (4.8) | 27,364 (9.0) | 0.17 |
| 22 Feb 2021 to 7 Mar 2021 | 3,539 (6.6) | 30,760 (11.4) | 0.17 | 1,491 (7.0) | 32,808 (10.8) | 0.13 |
| 8 Mar 2021 to 21 Mar 2021 | 5,134 (9.6) | 32,776 (12.1) | 0.08 | 2,790 (13.1) | 35,120 (11.6) | 0.05 |
| 22 Mar 2021 to 4 Apr 2021 | 8,338 (15.7) | 35,910 (13.3) | 0.07 | 4,814 (22.6) | 39,434 (13.0) | 0.25 |
| 5 Apr 2021 to 19 Apr 2021 | 13,252 (24.9) | 41,045 (15.2) | 0.24 | 8,526 (40.1) | 45,771 (15.1) | 0.58 |
| Number of tests in previous 3 months |  |  |  |  |  |  |
| 0 | 43,713 (82.1) | 189,786 (70.1) | 0.28 | 11,588 (54.5) | 221,911 (73.3) | 0.40 |
| 1 | 7,151 (13.4) | 54,827 (20.2) | 0.18 | 4,338 (20.4) | 57,640 (19.0) | 0.03 |
| ≥2 | 2,406 (4.5) | 26,150 (9.7) | 0.20 | 5,346 (25.1) | 23,210 (7.7) | 0.49 |
| Any comorbidity<sup>d</sup> | 23,212 (43.6) | 127,974 (47.3) | 0.07 | 12,218 (57.4) | 138,968 (45.9) | 0.23 |
| Receipt of 2019-2020 and/or 2020-2021<br>influenza vaccination | 13,751 (25.8) | 89,395 (33.0) | 0.16 | 9,587 (45.1) | 93,559 (30.9) | 0.30 |
| Household income quintile<sup>c, e</sup> |  |  |  |  |  |  |
| 1 (lowest) | 11,878 (22.3) | 47,944 (17.7) | 0.11 | 3,750 (17.6) | 56,072 (18.5) | 0.02 |
| 2 | 11,154 (20.9) | 51,470 (19.0) | 0.05 | 4,146 (19.5) | 58,478 (19.3) | 0.00 |
| 3 | 11,477 (21.5) | 52,628 (19.4) | 0.05 | 4,233 (19.9) | 59,872 (19.8) | 0.00 |
| 4 | 10,146 (19.0) | 56,676 (20.9) | 0.05 | 4,513 (21.2) | 62,309 (20.6) | 0.02 |
| 5 (highest) | 8,359 (15.7) | 60,774 (22.4) | 0.17 | 4,540 (21.3) | 64,593 (21.3) | 0.00 |
| Essential workers quintile<sup>c, f</sup> |  |  |  |  |  |  |
| 1 (0%–32.5%) | 6,440 (12.1) | 50,664 (18.7) | 0.18 | 3,917 (18.4) | 53,187 (17.6) | 0.02 |
| 2 (32.5%–42.3%) | 11,225 (21.1) | 60,040 (22.2) | 0.03 | 4,664 (21.9) | 66,601 (22.0) | 0.00 |
| 3 (42.3%–49.8%) | 11,106 (20.8) | 56,108 (20.7) | 0.00 | 4,468 (21.0) | 62,746 (20.7) | 0.01 |
| 4 (50.0%–57.5%) | 11,576 (21.7) | 52,849 (19.5) | 0.05 | 4,211 (19.8) | 60,214 (19.9) | 0.00 |
| 5 (57.5%–100%) | 12,519 (23.5) | 49,067 (18.1) | 0.13 | 3,859 (18.1) | 57,727 (19.1) | 0.02 |
| Persons per dwelling quintile<sup>c, g</sup> |  |  |  |  |  |  |
| 1 (0–2.1) | 5,781 (10.9) | 51,852 (19.2) | 0.23 | 4,277 (20.1) | 53,356 (17.6) | 0.06 |
| 2 (2.2–2.4) | 6,641 (12.5) | 52,326 (19.3) | 0.19 | 4,219 (19.8) | 54,748 (18.1) | 0.04 |
| 3 (2.5–2.6) | 5,633 (10.6) | 37,229 (13.7) | 0.10 | 3,020 (14.2) | 39,842 (13.2) | 0.03 |
| 4 (2.7–3.0) | 12,967 (24.3) | 63,774 (23.6) | 0.02 | 4,874 (22.9) | 71,867 (23.7) | 0.02 |
| 5 (3.1–5.7) | 21,833 (41.0) | 63,459 (23.4) | 0.38 | 4,709 (22.1) | 80,583 (26.6) | 0.10 |
| Self-identified visible minority quintile<sup>c, h</sup> |  |  |  |  |  |  |
| 1 (0.0%–2.2%) | 4,437 (8.3) | 51,919 (19.2) | 0.32 | 4,133 (19.4) | 52,223 (17.2) | 0.06 |
| 2 (2.2%–7.5%) | 5,752 (10.8) | 55,124 (20.4) | 0.27 | 4,592 (21.6) | 56,284 (18.6) | 0.07 |
| 3 (7.5%–18.7%) | 7,223 (13.6) | 51,122 (18.9) | 0.14 | 3,982 (18.7) | 54,363 (18.0) | 0.02 |
| 4 (18.7%–43.5%) | 10,718 (20.1) | 53,691 (19.8) | 0.01 | 3,974 (18.7) | 60,435 (20.0) | 0.03 |
| 5 (43.5%–100%) | 24,736 (46.4) | 56,876 (21.0) | 0.56 | 4,438 (20.9) | 77,174 (25.5) | 0.11 |

<sup>a</sup>Proportion reported, unless stated otherwise.
<sup>b</sup>Standardized differences of >0.10 are considered clinically relevant.
<sup>c</sup>The sum of counts does not equal the column total because of individuals with missing information (<1.0%) for this characteristic.
<sup>d</sup>Comorbidities include chronic respiratory diseases, chronic heart diseases, hypertension, diabetes, immunocompromising conditions due to underlying diseases or therapy, autoimmune diseases, chronic kidney disease, advanced liver disease, dementia/frailty and history of stroke or transient ischemic attack.
<sup>e</sup>Household income quintile has variable cut-off values in each city/Census area to account for cost of living. A dissemination area (DA) being in quintile 1 means it is among the lowest 20% of DAs in its city by income.
<sup>f</sup>Percentage of people in the area working in the following occupations: sales and service occupations; trades, transport and equipment operators and related occupations; natural resources, agriculture, and related production occupations; and occupations in manufacturing and utilities. Census counts for people are randomly rounded up or down to the nearest number divisible by 5, which causes some minor imprecision.
<sup>g</sup>Range of persons per dwelling.
<sup>h</sup>Percentage of people in the area who self-identified as a visible minority. Census counts for people are randomly rounded up or down to the nearest number divisible by 5, which causes some minor imprecision.

Against symptomatic infection, adjusted VE ≥14 days after receiving only 1 dose was 60% (95%CI, 57–64%). This increased from 48% (95%CI, 41–54%) at 14–20 days to a plateau of 71% (95%CI, 63–78%) at 35–41 days (**Figure 2, eTable 5**). We observed a 16% increase in risk of symptomatic infection 7-13 days after a first dose (VE -16%; 95%CI, -26% to -6%), but no increase 0-6 days after a first dose. VE ≥7 days after receiving 2 doses was 91% (95%CI, 89– 93%). Against severe outcomes of hospitalization or death, VE ≥14 days after receiving 1 dose was 70% (95%CI, 60–77%), increasing from 62% (95%CI, 44–75%) at 14–20 days to 91% (95%CI, 73–97%) at ≥35 days, whereas VE ≥7 days after receiving 2 doses was 98% (95%CI, 88–100%) (**Figure 3, eTable 5)**.

**Figure 2.**
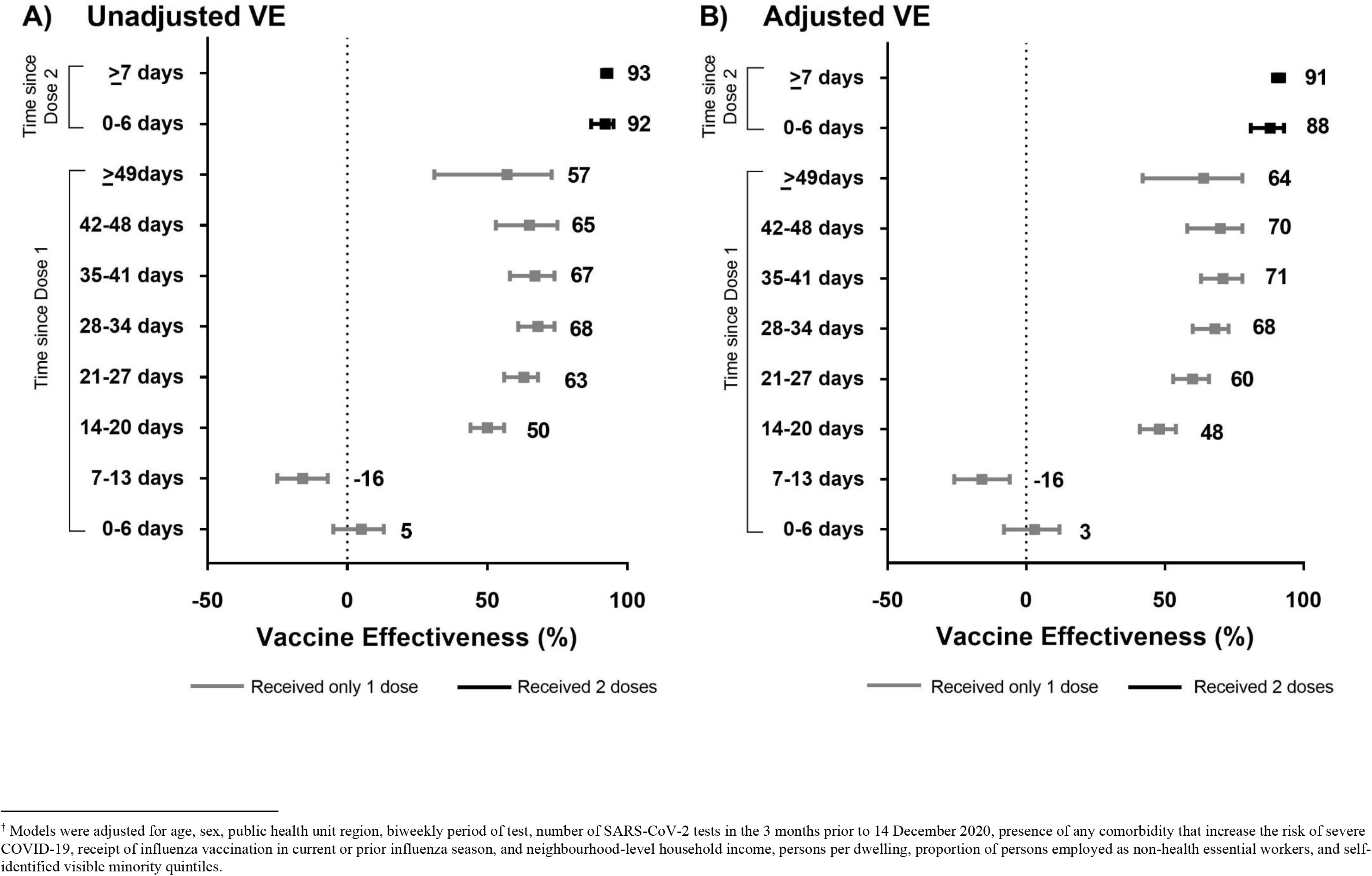
Unadjusted (panel A) and adjusted† (panel B) vaccine effectiveness estimates of COVID-19 mRNA vaccines (BNT162b2, mRNA-1273) against laboratory-confirmed symptomatic SARS-CoV-2 infection by various intervals, between 14 December 2020 and 19 April 2021 in Ontario, Canada.

**Figure 3.**
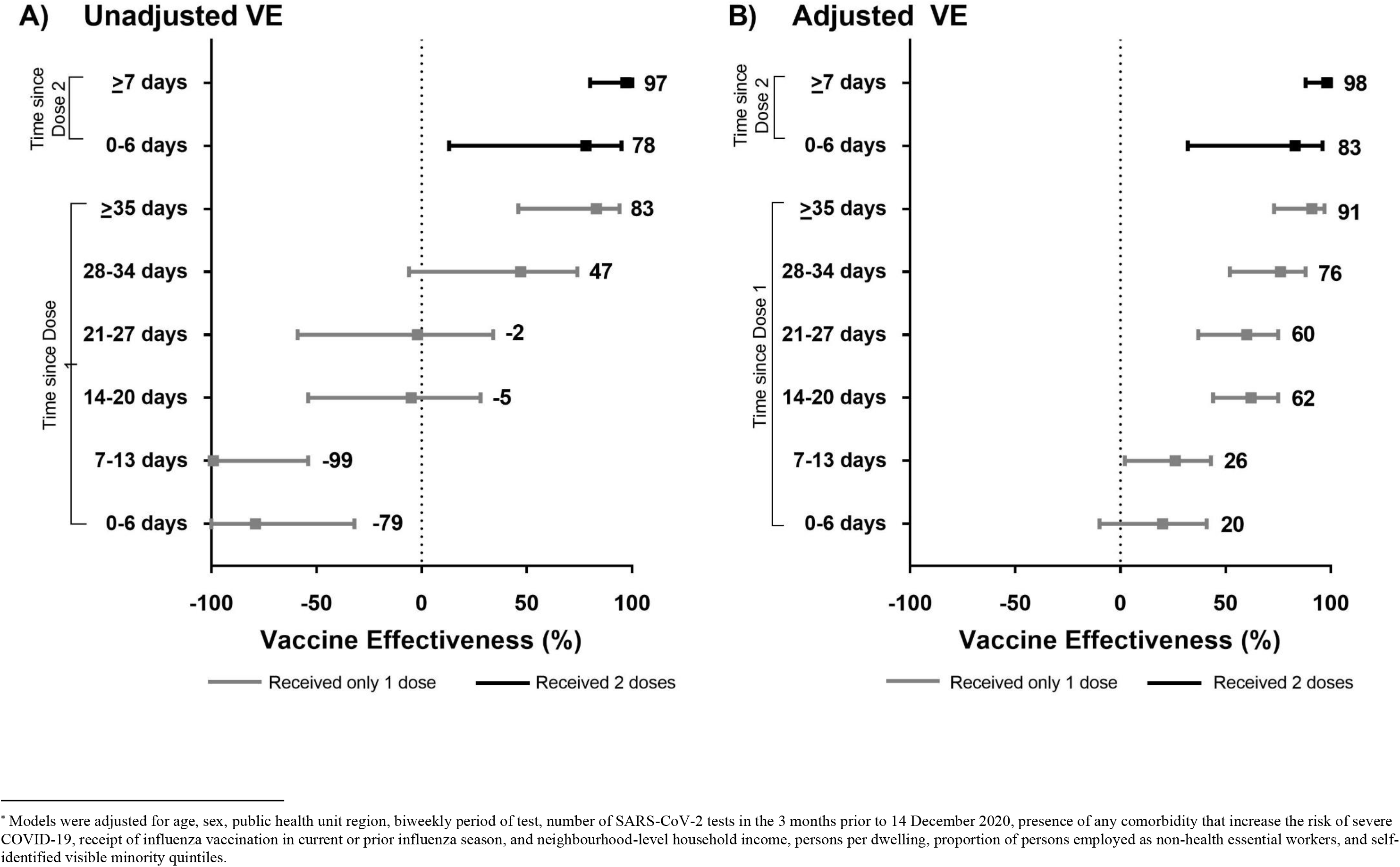
Unadjusted (panel A) and adjusted* (panel B) vaccine effectiveness estimates of COVID-19 mRNA vaccines (BNT162b2, mRNA-1273) against severe outcomes (hospitalization or death) associated with laboratory-confirmed symptomatic SARS-CoV-2 infection by various intervals, between 14 December 2020 and 19 April 2021 in Ontario, Canada.

In subgroup analyses of VE against symptomatic infection, we observed higher VE ≥14 days after receiving only 1 dose with mRNA-1273 than BNT162b2 (which was consistent across all age groups), for younger adults than adults aged ≥70 years, for individuals with no comorbidities than for those with comorbidities, and against the earlier variant and Alpha than Beta or Gamma (though 95% confidence intervals for VE for variants overlapped) (**Figure 4a, eTable 6**). However, VE estimates ≥7 days after receiving 2 doses were high (all ≥88%) and comparable across all subgroups, including against E484K-positive variants. Against severe outcomes, we observed higher VE ≥14 days after receiving 1 dose for younger adults aged 16-39 years, but VE estimates after receiving 2 doses were mostly similar across subgroups (**Figure 4b, eTable 7**).

**Figure 4.**
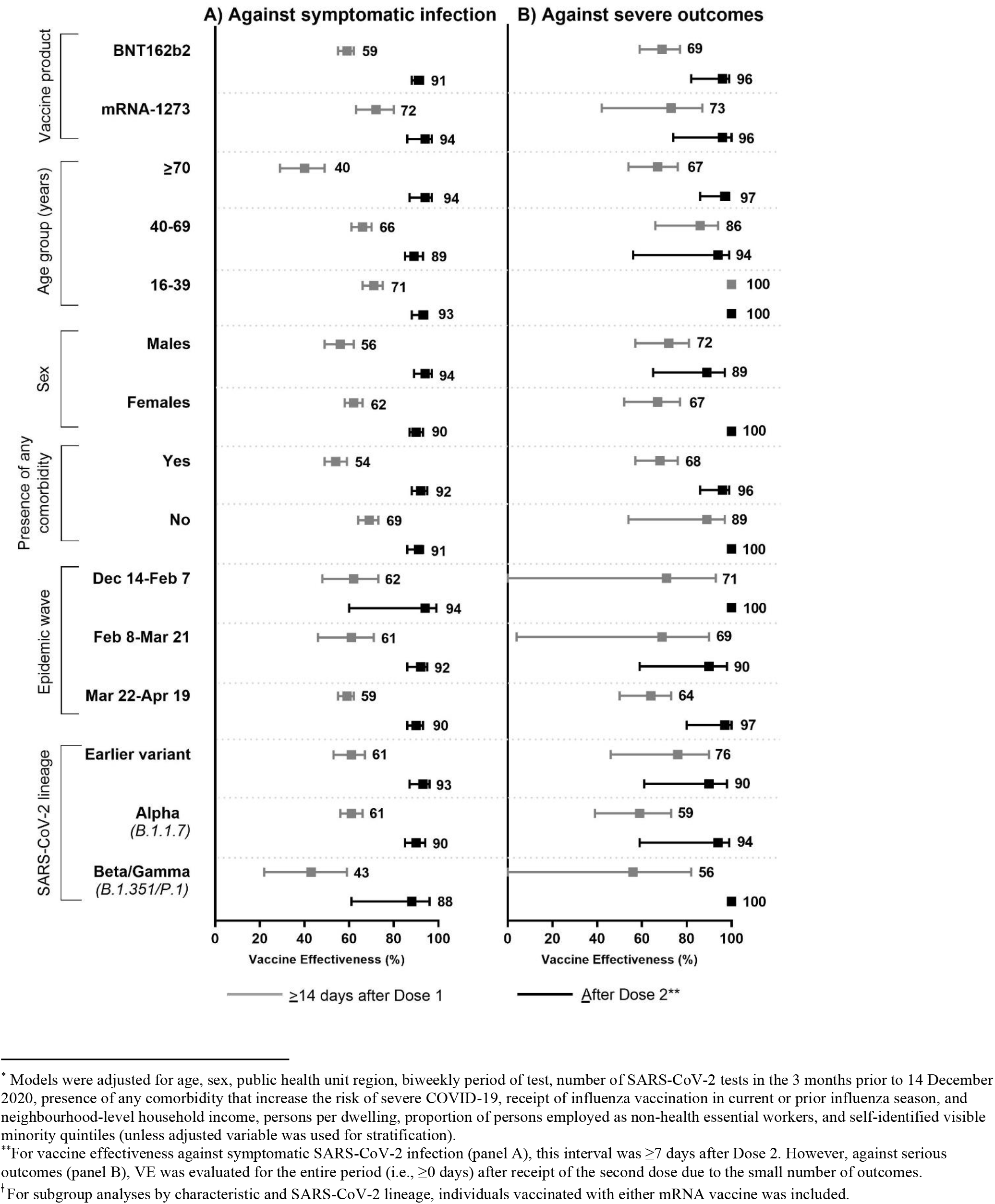
Adjusted* vaccine effectiveness estimates ≥14 days after Dose 1 (for individuals who received only 1 dose) and ≥0 days after Dose 2 by various factors, including vaccine product, patient characteristics, epidemic wave, and SARS-CoV-2 lineage against laboratory-confirmed symptomatic SARS-CoV-2 infection (panel A) and severe outcomes (hospitalization or death) (panel B) between 14 December 2020 and 19 April 2021.

Among adults ≥70 years, VE against symptomatic infection after 1 dose was 64% (95%CI 46– 76%) at 28–34 days and 85% (95%CI 38–97%) at 42–48 days, whereas comparable VE estimates were achieved sooner after 1 dose for younger adults (**Figure 5, eTable 8**). Furthermore, VE against severe outcomes was similar at ≥35 days after 1 dose (93%; 95%CI, 71–98%) as after receiving 2 doses (97%; 95%CI, 86–99%).

**Figure 5.**
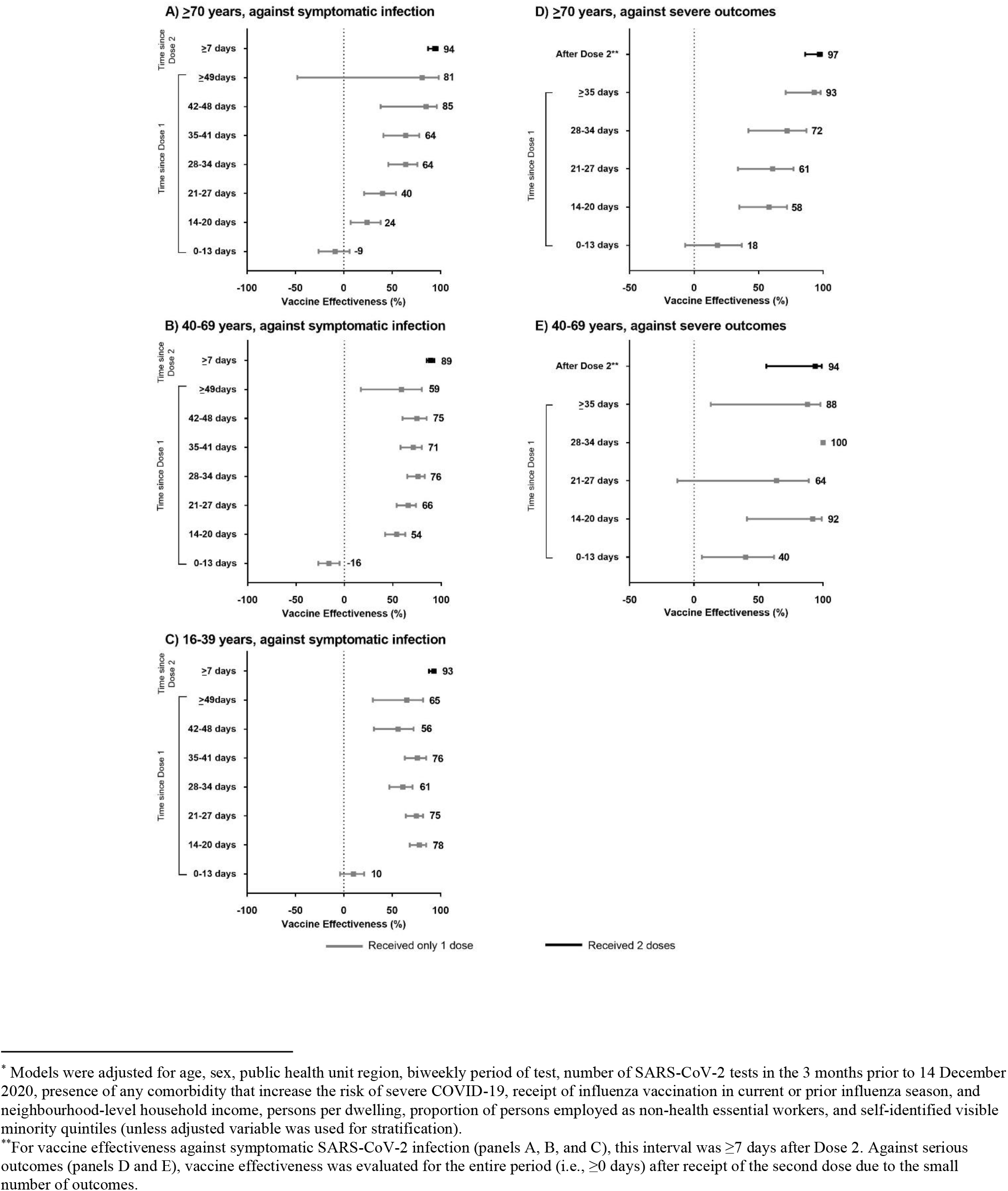
Adjusted* vaccine effectiveness estimates ≥14 days after Dose 1 and ≥0 days after Dose 2 in community-dwelling adults in Ontario, Canada against laboratory-confirmed symptomatic SARS-CoV-2 infection for adults aged ≥70 years (panel A), 40-69 years (panel B), and 16-39 years (panel C) and severe outcomes (hospitalization or death) for adults aged ≥70 years (panel D) and 40-69 years (panel E), between 14 December 2020 and 19 April 2021.

Lastly, in the sensitivity analysis that restricted the analysis to vaccinated individuals and treated those vaccinated 0-13 days prior to the index date as the reference group, VE estimates against symptomatic infection were similar to the main analyses (**eTable 5)**. However, for severe outcomes, the estimates differed for earlier vaccination intervals.

## DISCUSSION

Using the test-negative design, which mitigates selection bias due to differences in health-seeking behaviour between vaccinated and unvaccinated individuals, we estimated very high (>90%) vaccine effectiveness of mRNA vaccines BNT162b2 and mRNA-1273 against symptomatic SARS-CoV-2 infection with full vaccination (i.e., ≥7 days after receipt of a second dose), and moderate (∼50-70%) VE with partial vaccination (i.e., ≥14 days after receipt of only a first dose). Estimates for both full and partial vaccination were approximately 10 percentage points higher against hospitalization or death than symptomatic infection. VE generally increased over time after a first dose, however we also noted a slightly increased risk of symptomatic infection on days 7-13 after a first dose, compared to no vaccination. In subgroup analyses, we observed lower VE against symptomatic infection in adults aged ≥70 years and individuals with comorbidities, but a higher VE after the first dose for mRNA-1273 recipients than BNT162b2 recipients. However, VE was consistently high across subgroups for fully vaccinated individuals, and also for older adults after longer intervals following receipt of a first dose.

Our findings for fully vaccinated individuals are comparable with clinical trial efficacy estimates and other real-world effectiveness estimates reported in a range of settings.[16,18–31] Existing evidence estimating one-dose effectiveness of mRNA vaccines from observational studies is heterogeneous,[16,27,30–32] with estimates for symptomatic infection ranging from 57% (95%CI, 50–63%)[16] to 72% (95%CI, 58–86%)[31] and post-hoc calculations from efficacy trials approximately 90%.[33,34] There is similar heterogeneity among one-dose effectiveness estimates in older adults,[25,32,35] with estimates generally lower for older adults after the first dose,[16,32] and increasing with time. Our analysis identified an effectiveness against symptomatic infection of 63% (95%CI, 40–72%) ≥49 days after only the first dose, in keeping with several other studies reporting one-dose effectiveness.[16,30] In addition, we found that one dose of the mRNA-1273 vaccine had a significantly higher VE against symptomatic infection than BNT162b2. Differences in characteristics between BNT162b2 and mRNA-1273 recipients might explain this finding, but similar results were also found in another Canadian province.[36] However findings were inconsistent in other studies that compared the effectiveness between products after one dose; some found a trend towards higher VE against infection in mRNA-1273 recipients,[37,38] whereas others found no difference.[39,40] However for each, their study populations were more homogeneous than ours (e.g., adults aged ≤40 years, healthcare workers, veterans). Our analysis also reflects extant evidence that effectiveness increases to very high levels after the second dose, even in older adults against symptomatic infection[16] and COVID-19-associated hospitalizations.[19,23] Lastly, our finding that receipt of 2 doses of mRNA vaccines was not associated with appreciable vaccine escape by lineage Alpha or E484K-positive variants (i.e., Beta and Gamma) is notable.

In our study, we found an increased risk of infection 7-13 days after receiving the vaccine. An increased risk of SARS-CoV-2 infection up to 14 days after the first dose was also found in other studies.[25,31,41,42] This could be due to an increase in SARS-CoV-2 exposures after vaccination. Individuals may assume protection immediately following vaccination and engage in higher risk behaviours before a sufficient immune response has developed. Indeed, approximately 20% of the US public believe that protection is conferred either immediately or 1-2 weeks after the first dose.[43] Future studies should examine the potential role of behavioural changes post first dose of COVID-19 vaccines. This finding could also be due a higher baseline risk of infection among those who were initially prioritized to receive the vaccine, which may not have been adequately controlled for in our models. However, a VE estimate of close to 0% 0-6 days after a first dose provides a level of validation that differences between vaccinated and unvaccinated individuals were accounted for.

Our study had some limitations. First, our study sample was limited to those with COVID-19 symptoms recorded in OLIS, which decreased our potential sample size considerably, from 2,171,449 individuals who were tested for SARS-CoV-2 to 324,033 individuals who had relevant COVID-19 symptoms recorded in OLIS. Not all laboratories in Ontario currently have the information technology infrastructure to submit symptom information (or documentation of asymptomatic testing) recorded on the SARS-CoV-2 laboratory requisition into OLIS. Thus, the generalizability of our findings to the broader population is uncertain and we could not evaluate VE against asymptomatic infection. However, the percent positivity of our study sample (53,270/324,033=16.4%) was not dramatically different from the percent positivity observed in Ontario during the study period (281,261/2,171,449=13.0%), and we would expect positivity to be higher for symptomatic individuals than asymptomatic individuals. We also acknowledge that those who had no symptom information recorded in OLIS may have been symptomatic at the time of testing and those recorded as asymptomatic may have subsequently become symptomatic. In addition, COVID-19 vaccination status may also be collected on the laboratory requisition. This may bias the true VE estimate, depending on whether symptoms were more likely to be documented on requisition forms for vaccinated individuals who ultimately test positive for SARS-CoV-2 (bias VE towards the null) or have symptoms less likely recorded (bias VE away from the null). Traditional test-negative design studies collect vaccination status among all individuals with symptoms consistent with the pathogen under study to minimize this selection bias. However, the congruence of our findings for fully vaccinated individuals with extant studies provides some reassurance that any under- or overestimation of VE is likely to be small. Second, because symptom onset date is largely unavailable in OLIS and CCM only has information on test-positive cases, we used specimen collection date as the index date. This may have led to classifying some vaccinated individuals into an incorrect dose-to-index date interval because their symptom onset would have been several days prior to getting tested. The impact to VE estimates for earlier intervals if using the specimen collection date depends on the test results for vaccinated individuals (e.g., VE for earlier intervals would be *overestimated* if there were more vaccinated test-positive cases and *underestimated* if there were more vaccinated test-negative controls) or whether the lag between symptom onset and specimen collection dates resulted in misclassification of vaccinated individuals (i.e., false negatives), which would *overestimate* VE for earlier intervals. Furthermore, we could not limit the study population to individuals tested within 10 days of symptom onset, a commonly used inclusion criterion for test-negative studies. Prolonging the interval between symptom onset and testing increases the likelihood of false-negative cases, which lowers VE estimates. However, 89% of cases with both symptom onset and specimen collection date documented in CCM (not the source of symptom data for this study) were tested within 10 days of symptom onset. Third, our results may have been impacted by outcome misclassification of severe outcomes due to unlinked case records and incomplete capture of severe outcomes in CCM, and delays in identifying hospitalizations in DAD (which are dependent on individuals being discharged) and deaths in RPDB. The direction of bias to VE estimates depends on whether data completeness and lags are differential between vaccinated and unvaccinated test-positive cases (e.g., if vaccination status ascertained during the case management process influenced the degree of data collection [e.g., if more complete among vaccinated cases, VE would be biased towards the null], or if unvaccinated cases have a prolonged hospitalization because of more severe course of illness, their hospitalization record would not be available for analysis and VE would be biased towards the null). Fourth, some of our covariates may be subject to measurement error. We used frequency of previous SARS-CoV-2 tests as a proxy to identify individuals at higher risk of exposure (and increased likelihood to be targeted for early vaccination). However, we did not include point-of-care tests because they are incompletely captured in OLIS. Furthermore, since access to testing is variable, we might not have adequately controlled for this concept. Finally, we may not have adequately accounted for confounding bias with the covariates that were available in the study databases, especially against severe outcomes.

Our findings suggest that older individuals and those with comorbidities may benefit from risk-based recommendations to minimize second-dose delays. However, rising protection against severe outcomes – arguably the more important outcome – with increasing time after a first dose provides support for delaying the second dose in settings that face vaccine supply constraints. Mathematical modelling could be conducted to demonstrate how, particularly for jurisdictions with limited vaccine supply, vaccines should be distributed to maximize population protection (e.g., the relative benefits of providing second doses earlier to older populations versus providing more first doses to younger populations who respond better to a single dose and leading to more rapid achievement of herd immunity by maximizing coverage with 1 dose). Since VE against symptomatic infection after 1 dose is only moderate, and among older adults appears to be modest even at 14–20 days, individuals need to be informed that besides the absence of benefit during the first 2 weeks (and likely longer for older adults) after a first dose, they should continue to adhere to recommended public health measures, such as mask-wearing, physical distancing, and avoidance of social gatherings.

## Data Availability

The dataset from this study is held securely in coded form at ICES. While legal data sharing agreements between ICES and data providers (e.g., healthcare organizations and government) prohibit ICES from making the dataset publicly available, access may be granted to those who meet pre-specified criteria for confidential access, available at www.ices.on.ca/DAS. The full dataset creation plan and underlying analytic code are available from the authors upon request, understanding that the computer programs may rely upon coding templates or macros that are unique to ICES and are therefore either inaccessible or may require modification.

## Contributors

HC and JCK designed and oversaw the study. SH and HC obtained the data and conducted all analyses (data set and variable creation and statistical modelling). BC contributed to data analyses and data preparation for the symptomatic data set. SN, MES, HC, and JCK drafted the manuscript. All authors contributed to the analysis plan, interpreted the results, critically reviewed and edited the manuscript, approved the final version, and agreed to be accountable for all aspects of the work.

## Ethics approval

ICES is a prescribed entity under Ontario’s Personal Health Information Protection Act (PHIPA). Section 45 of PHIPA authorizes ICES to collect personal health information, without consent, for the purpose of analysis or compiling statistical information with respect to the management of, evaluation or monitoring of, the allocation of resources to or planning for all or part of the health system. Projects that use data collected by ICES under section 45 of PHIPA, and use no other data, are exempt from REB review. The use of the data in this project is authorized under section 45 and approved by ICES’ Privacy and Legal Office.

## Dissemination statement

As this research involves information drawn from the entire population of Ontario, it is not feasible to report these results to each individual Ontarian. However, the results from this manuscript will be made publicly available through a pre-print server and through dissemination by ICES, news media, and other means.

## Transparency declaration

JCK affirms that the manuscript is an honest, accurate, and transparent account of the study being reported; that no important aspects of the study have been omitted; and that any discrepancies from the study as planned have been explained.

## Acknowledgments

We would like to acknowledge Public Health Ontario for access to case-level data from CCM and COVID-19 laboratory data, as well as assistance with data interpretation. We also thank the staff of Ontario’s public health units who are responsible for COVID-19 case and contact management and data collection within CCM. We thank IQVIA Solutions Canada Inc. for use of their Drug Information Database. The authors are grateful to the Ontario residents without whom this research would be impossible.

## Disclaimers

This study was supported by ICES, which is funded by an annual grant from the Ontario Ministry of Health (MOH) and the Ministry of Long-Term Care (MLTC). This study was supported by the Ontario Health Data Platform (OHDP), a Province of Ontario initiative to support Ontario’s ongoing response to COVID-19 and its related impacts. The study sponsors did not participate in the design and conduct of the study; collection, management, analysis and interpretation of the data; preparation, review or approval of the manuscript; or the decision to submit the manuscript for publication. Parts of this material are based on data and/or information compiled and provided by the Canadian Institute for Health Information (CIHI) and by Cancer Care Ontario (CCO). However, the analyses, conclusions, opinions and statement expressed herein are solely those of the authors, and do not reflect those of the funding or data sources; no endorsement by ICES, MOH, MLTC, OHDP, its partners, the Province of Ontario, CIHI or CCO is intended or should be inferred.

## Licence for Publication

The Corresponding Author has the right to grant on behalf of all authors and does grant on behalf of all authors, an exclusive licence (or non exclusive for government employees) on a worldwide basis to the BMJ Publishing Group Ltd to permit this article (if accepted) to be published in BMJ and any other BMJPGL products and sublicences such use and exploit all subsidiary rights, as set out in our licence (http://group.bmj.com/products/journals/instructions-for-authors/licence-forms).

