## Supplementary for "Effectiveness of BNT162b2 and mRNA-1273 COVID-19 vaccines against symptomatic SARS-CoV-2 infection and severe COVID-19 outcomes in Ontario, Canada: a test-negative design study"

**Supplementary Material:**

**Supplementary Methods:** Determination of symptom status at the time of SARS-CoV-2 testing

**eTable 1:** Definitions of covariates used in the analysis.

**eTable 2:** Comparison of unadjusted vaccine effectiveness estimates and VE estimates adjusted for a single covariate to evaluate the impact of potential confounders.

**eFigure 1:** Distribution of mRNA vaccine product (BNT162b2, mRNA-1273) among individuals included in the study cohort and all adults aged ≥16 years between 14 December 2020 and 19 April 2021 in Ontario, Canada.

**eTable 3:** Characteristics of individuals with symptoms consistent with COVID-19 recorded in the Ontario Laboratories Information System (OLIS) at the time of testing for SARS-CoV-2, individuals recorded in OLIS as asymptomatic, and individuals who did not have symptoms consistent with COVID-19 or had no symptom information recorded in OLIS between 14 December 2020 and 19 April 2021 in Ontario, Canada.

**eTable 4:** Characteristics of symptomatic individuals tested for SARS-CoV-2 according to vaccine product and vaccination status between 14 December 2020 and 19 April 2021 in Ontario, Canada.

**eTable 5:** Unadjusted and adjusted vaccine effectiveness estimates of COVID-19 mRNA vaccines against symptomatic SARS-CoV-2 infection and severe outcomes (hospitalization or death) by various intervals, between 14 December 2020 and 19 April 2021 in Ontario, Canada.

**eTable 6:** Unadjusted and adjusted vaccine effectiveness estimates against symptomatic COVID-19 infection after Dose 1 (for individuals who received only 1 dose) and Dose 2, by various factors, including vaccine product, patient characteristics, epidemic wave, and SARS-CoV-2 lineage between 14 December 2020 and 19 April 2021 in Ontario, Canada.

**eTable 7:** Unadjusted and adjusted vaccine effectiveness estimates after Dose 1 (for individuals who received only 1 dose) and Dose 2, by various factors, including vaccine product, patient characteristics, epidemic wave, and SARS-CoV-2 lineage against severe outcomes (hospitalization or death) of symptomatic SARS-CoV-2 infection between 14 December 2020 and 19 April 2021 in Ontario, Canada.

**eTable 8:** Unadjusted and adjusted vaccine effectiveness estimates against symptomatic SARS-CoV-2 infection and severe outcomes (hospitalization or death), stratified by age group, between 14 December 2020 and 19 April 2021 in Ontario, Canada.

CONFIDENTIAL

**Supplementary Methods.** Determination of symptom status at the time of SARS-CoV-2 testing

The Ontario Laboratories Information System (OLIS) contains open fields (specifically, using the Patient Note Clinical Information field or reporting under the observation code XON13543-4 [Patient symptoms]) that records whether individuals tested for SARS-CoV-2 presented with symptoms at the time of the test. These character-based fields were originally delimited by commas, slashes, semicolons, or ampersands. Character text-strings were parsed and aggregated.

One of the authors, BC, identified symptom classifications available in OLIS as of April 19, 2021 that were likely to be due to COVID-19 and issued a list of symptoms to be adjudicated with JK. Low-frequency terms (appearing <25 times throughout) were excluded. Values listed in the symptoms fields were classified by JK and BC into “symptomatic” and “asymptomatic”. We purposely chose a broadly inclusive definition to capture all potentially relevant COVID-19 symptoms, including atypical symptoms and chronic conditions, based on our scientific and medical understanding of COVID-19-related symptoms as of April 19, 2021.[1]

Terms determined to be indicative of COVID-19 symptoms (classified as ‘symptomatic’) are listed below. In addition to this list, we used SYMPTOMATIC (or partial spellings thereof) or mention of symptom onset.

0 TASTE, 37.8, 37.9, 38.0, 38.1, 38.2, 38.3, 38.4, 38.5, 38.6, 38.7, 38.8, 38.9, 39.0, 39.1, 39.2, 39.3, 39.4, 39.5, 100, 101, 102, 38, 39, ABD, ABD PAIN, ABD. PAIN, ABD.PAIN, ABDO CRAMPS, ABDO PAIN, ABDOMINAL, ABDOMINAL PAIN, ACHE, ACHES, ACHES AND PAINS, ACHES HEADACHE, ACHEY, ACHINESS, ACHY, ALLERGIES, ANOREXIA, AP, APPETITE, ARTHRALGIA, ASTHMA, BACK ACHE, BACK PAIN, BACKACHE, BLOATING, BLOODY STOOLS, BODY ACHE, BODY ACHES, BODY ACHES FATIGUE, BODY ACHES HEADACHE, BODY CHILLS, BODY PAIN, BODY PAINS, BODYACHE, BODYACHES, CHANGE IN TASTE, CHEST, CHEST BURNING, CHEST CONGESTION, CHEST DISCOMFORT, CHEST HEAVINESS, CHEST HEAVY, CHEST PAIN, CHEST PAINS, CHEST PRESSURE, CHEST TIGHT, CHEST TIGHTNESS, CHESTPAIN, CHF, CHI, CHILL, CHILLS, CHILLS BODY ACHES, CHILLS FATIGUE, CHILLS HEADACHE, CHRONIC COUGH, CLEARING THROAT, COLD, COLD LIKE SYMPTOMS, COLD SWEATS, COLD SYMPTOMS, COLD-LIKE SYMPTOMS, CONFUSION, CONG, CONGE, CONGES, CONGEST, CONGESTED, CONGESTED COUGH, CONGESTED HEADACHE, CONGESTED NOSE, CONGESTED RUNNY NOSE, CONGESTI, CONGESTIO, CONGESTION, CONGESTION FATIGUE, CONGESTION HEADACHE, CONGESTION RUNNY NOSE, CONGESTIONS, CONJESTION, CONJUNCTIVITIS, CONSTIPATION, COPD, COUGH, COUGH ONSET 20, COUGHING, CP, CRAMPING, CRAMPS, DECREASE APPETITE, DECREASED APPETITE, DECREASED TASTE, DEHYDRATION, DELIRIUM, DIA, DIAHERRA, DIAHREA, DIAHRREA, DIAR, DIARHEA, DIARHHEA, DIARR, DIARRH, DIARRHE, DIARRHEA, DIARRHEA BLOODY, DIARRHEA FATIGUE, DIARRHEA HEADACHE, DIARRHEA NAUSEA,

CONFIDENTIAL

DIARRHEA VOMITING, DIARRHEA WATERY, DIARRHOEA, DIFFICULTY BREATHING, DIFFICULTY SWALLOWING, DIRRHEA, DISCOMFORT, DIZZINESS, DIZZY, DRY COUGH, DRY THROAT, DYSPEPSIA, DYSPNEA, DYSURIA, EAR ACHE, EAR INFECTION, EAR PAIN, EARACHE, EMESIS, ENCEPHALITIS, EPIGASTRIC PAIN, EXHAUSTED, EXHAUSTION, EXTREME FATIGUE, FAT, FATI, FATIG, FATIGU, FATIGUE, FATIGUE AND HEADACHE, FATIGUE BODY ACHES, FATIGUE CHILLS, FATIGUE CONGESTION, FATIGUE DIARRHEA, FATIGUE HEADACHE, FATIGUE RUNNY NOSE, FATIGUE., FATIGUED, FATIQUE, FEBRILE, FEELING UNWELL, FEVER, FEVERISH, FLANK PAIN, FLU, FLU LIKE SYMPTOMS, GASTRITIS, GASTRO, GASTRO SYMPTOMS, GASTROENTERITIS, GEN WEAKNESS, GENERAL MALAISE, GENERAL WEAKNESS, GENERALLY UNWELL, GERD, GI, GI BLEEDING, GI ISSUE, GI ISSUES, GI SYMPTOMS, GI UPSET, HA, HALLUCINATIONS, HEA, HEAD, HEAD ACHE, HEAD ACHES, HEAD COLD, HEAD CONGESTION, HEADA, HEADAC, HEADACH, HEADACHE, HEADACHE (ONLY), HEADACHE AND BODY ACHES, HEADACHE AND CHILLS, HEADACHE AND CONGESTION, HEADACHE AND FATIGUE, HEADACHE AND NAUSEA, HEADACHE AND RUNNY NOSE, HEADACHE BODY ACHE, HEADACHE BODY ACHES, HEADACHE CHILLS, HEADACHE CONGESTED, HEADACHE CONGESTION, HEADACHE DIARRHEA, HEADACHE FATIGUE, HEADACHE NASAL CONGESTION, HEADACHE NAUSEA, HEADACHE RUNNY NOSE, HEADACHE SOB, HEADACHE STOMACH ACHE, HEADACHE STUFFY NOSE, HEADACHE TIRED, HEADACHE UPSET STOMACH, HEADACHE VOMITING, HEADACHE., HEADACHE. FATIGUE, HEADACHE.VSS, HEADACHES, HEADAHCE, HEADCHE, HEART FAILURE, HEARTBURN, HEAVINESS IN CHEST, HEAVY CHEST, HEMATURIA, HIA, HOARSE, HOARSE VOICE, HOT, HOT FLASHES, ITCHY THROAT, JAUNDICE, JOINT PAIN, LACK OF APPETITE, LETHARGIC, LETHARGY, LIGHT HEADED, LIGHTHEADED, LOA, LOOSE BM, LOOSE STOOL, LOOSE STOOLS, LOS, LOSS APPETITE, LOSS OF APPETITE, LOSS OF SMELL, LOSS OF SMELL AND TASTE, LOSS OF TASTE, LOSS OF TASTE AND SMELL, LOSS OF VOICE, LOSS SMELL, LOSS TASTE, LOSS TASTE AND SMELL, LOST OF TASTE, LOST TASTE, LOW APPETITE, LOW ENERGY, LOW GRADE FEVER, LOWER BACK PAIN, MACULOPAPULAR RASH, MALAISE, MENINGITIS, MIGRAINE, MIGRAINES, MILD HEADACHE, MUSCLE, MUSCLE ACHE, MUSCLE ACHES, MUSCLE PAIN, MUSCLE WEAKNESS, MUSCLEACHES, MUSCLES ACHES, MYALGIA, MYALGIAS, N\T\V, N+V, NAS.CONG, NASAL, NASAL CONG, NASAL CONG., NASAL CONGEST, NASAL CONGESTED, NASAL CONGESTI, NASAL CONGESTIO, NASAL CONGESTION, NASAL CONGESTION HEADACHE, NASAL DISCHARGE, NASAL DRAINAGE, NASAL DRIP, NASAL SYMPTOMS, NASUEA, NAU, NAUS, NAUSE, NAUSEA, NAUSEA AND HEADACHE, NAUSEA AND VOMITING, NAUSEA DIARRHEA, NAUSEA FATIGUE, NAUSEA HEADACHE, NAUSEA VOMITING, NAUSEA VOMITING DIARRHEA, NAUSEATED, NAUSEOUS, NECK PAIN, NEW SMELL, NIGHT SWEATS, NO APPETITE, NO ENERGY, NO SMELL, NO SMELL OR TASTE, NO TASTE, NO TASTE OR SMELL, NON-SPECIFIC SYMPTOM(S) - SURVEILLANCE, NOSE, NOSE CONGESTION, NSTEMI, PAIN, PAINS, PALPITATIONS, PHLEGM, PINK EYE, PLUGGED EARS, PND, PNEUMONIA, POST NASAL, POST NASAL DRIP, PRODUCTIVE COUGH, R.NOSE, RASH, RASH - NOT

CONFIDENTIAL

SPECIFIED, RASPY VOICE, RECENT FEVER, RESPIRATORY SYMPTOMS, RHINITIS, RHINNORHEA, RHINO, RHINORHEA, RHINORREA, RHINORRHEA, RUN NOSE, RUNNING NOSE, RUNNING NOSE.VSS, RUNNU NOSE, RUNNY, RUNNY NOSE, RUNNY N, RUNNY NO, RUNNY NOS, RUNNY NOSE, RUNNY NOSE AND CONGESTION, RUNNY NOSE AND HEADACHE, RUNNY NOSE AND SNEEZING, RUNNY NOSE CONGESTED, RUNNY NOSE CONGESTION, RUNNY NOSE DIARRHEA, RUNNY NOSE FATIGUE, RUNNY NOSE HEADACHE, RUNNY NOSE ONSET 20, RUNNY NOSE OR SNEEZING, RUNNY NOSE SNEEZING, RUNNY NOSE., RUNNY NOSR, RUNNYNOSE, RUNY NOSE, SCRATCHY THROAT, SEIZURE, SEPSIS, SHAKES, SHORT OF BREATH, SHORTNESS OF BREATH, SINUS, SINUS CONGESTED, SINUS CONGESTION, SINUS HEADACHE, SINUS INFECTION, SINUS ISSUES, SINUS PAIN, SINUS PRESSURE, SINUSES, SLUGGISH, SMELL, SNEEZ, SNEEZE, SNEEZING, SNEEZING RUNNY NOSE, SNEEZY, SNIFFLES, SOB, SOBOE, SORE, SORE BACK, SORE CHEST, SORE EARS, SORE MUSCLES, SORE NECK, SORE STOMACH, SORE THR, SORE THROAT, SORE THROAT ONSET 20, SORE THT, SORE TUMMY, SORETHROAT, SORETHT, STEMI, STHROAT, STIFF NECK, STOMACH, STOMACH ACHE, STOMACH ACHES, STOMACH CRAMPS, STOMACH PAIN, STOMACH PAINS, STOMACH UPSET, STOMACHACHE, STOMACHE ACHE, STROKE, STUFF NOSE, STUFFED NOSE, STUFFED UP, STUFFING NOSE, STUFFY, STUFFY NOSE, STUFFY NOSE HEADACHE, STUFFYNOSE, SWEAT, SWEATING, SWEATS, SWEATY, SWELLING, SWOLLEN GLANDS, SWOLLEN LYMPH NODES, SWOLLEN TONSILS, SX, SYNCOPE, TACHYCARDIA, TACHYPNEA, TASTE, TASTE DISORDER, TEMPERATURE, TEMPERATURE: 100, TEMPERATURE: 101, TEMPERATURE: 102, TEMPERATURE: 103, TEMPERATURE: 37.8, TEMPERATURE: 37.9, TEMPERATURE: 38, TEMPERATURE: 38.0, TEMPERATURE: 38.1, TEMPERATURE: 38.2, TEMPERATURE: 38.3, TEMPERATURE: 38.4, TEMPERATURE: 38.5, TEMPERATURE: 38.6, TEMPERATURE: 38.7, TEMPERATURE: 38.8, TEMPERATURE: 38.9, TEMPERATURE: 39, TEMPERATURE: 39.0, TEMPERATURE: 39.1, TEMPERATURE: 39.2, TEMPERATURE: 39.3, THROAT, TICKLE IN THROAT, TIGHT CHEST, TIGHTNESS, TIGHTNESS IN CHEST, TIRED, TIREDNESS, TROUBLE BREATHING, TUMMY ACHE, UNEXP FATIGUE, UNEXPLAINED FATIGUE, UNKNOWN COUGH, UNKNOWN FEVER, UNKNOWN PNEUMONIA, UNKNOWN SOB, UNKNOWN SORE THROAT, UNWELL, UPSET STOMACH, UPSET STOMACHE, URINARY TRACT INFECTION, VERTIGO, VESICULAR RASH, VOM, VOMIT, VOMITED, VOMITING, VOMITING AND DIARRHEA, VOMITING DIARRHEA, VOMITTED, VOMITTING, VOMMITING, VOMMITTING, WARM, WATERY EYES, WEAK, WEAKNESS, WEIGHT LOSS, WHEEZE, WHEEZING, WHEEZY, YES COUGH, YES FEVER, YES PNEUMONIA, YES SOB, YES SORE THROAT

Terms determined to be indicative of the absence of COVID-19 symptoms (classified as ‘asymptomatic’) were as follows: ASYMPTOMATIC (or partial spellings thereof). We also considered other terms that explicitly implied the negative presence of COVID-19 symptoms (e.g., NONE, NO PNEUMONIA, NO COUGH, NO FEVER, NO SORE THROAT, NO SYMPTOMS, N, NO, NO SOB, NIL). If both symptomatic and

CONFIDENTIAL

asymptomatic terms were presented in a record, the individual was classified as ‘symptomatic’.

Among all eligible individuals tested for SARS-CoV-2 during the study period (n=2,137,686), 39% had a test record that noted whether the individual was symptomatic for COVID-19 or asymptomatic at the time of testing. For the remainder, these fields were blank, contained irrelevant information (e.g., indication of test [e.g., pre-op], targeted patient population [e.g., health care worker, close contact]), or recorded symptoms not consistent with COVID-19 (e.g., anxiety, falls).

CONFIDENTIAL

**eTable 1:** List of covariates used in the analyses.

| Variable | Definition |
|---|---|
| Age | Age was determined from the Registered Persons Database. This variable was included *a priori* as hypothesized to be directly related to COVID-19 infection risk.[2] |
| Sex | Sex was determined from the Registered Persons Database. This variable was included *a priori* as hypothesized to be directly related to COVID-19 infection risk.[2] |
| Biweekly period of COVID-19 test | Based on the index date (i.e. specimen collection date, or date of severe outcome if before specimen collection date):<br><br>14 Dec 2020 to 27 Dec 2020<br>28 Dec 2020 to 10 Jan 2021<br>11 Jan 2021 to 24 Jan 2021<br>25 Jan 2021 to 7 Feb 2021<br>8 Feb 2021 to 21 Feb 2021<br>22 Feb 2021 to 7 Mar 2021<br>8 Mar 2021 to 21 Mar 2021<br>22 Mar 2021 to 4 Apr 2021<br>5 Apr 2021 to 19 Apr 2021 |
| Chronic heart disease | Individuals were defined as having "chronic heart disease" if they had congestive heart failure (CHF), ischemic heart disease, or atrial fibrillation. The definitions for these conditions are as follows:<br><br><u>CHF:</u>[3]<br>An ICES-derived CHF database was used to identify patients with CHF, based on 1 NACRS, DAD, SDS, or OHIP claim and a second claim (from either) in 1 year. The CHF database is limited to those aged 40 years or older. This variable was included a priori as hypothesized to be directly related to COVID-19 infection risk.[4]<br><br>OHIP: 428<br>DAD, SDS: ICD-9: 428, ICD-10: I500, I501, I509<br><br><u>Cardiac ischemic disease:</u>[5]<br>Any comorbidity in the past 5 years (DAD, any diagnosis field) or history of procedure in past 20 years (DAD, SDS), of the following:<br><br>Comorbidity (DAD, any diagnosis in the past 5 years):<br>Angina: ICD-10: I20<br><br>Chronic Ischemic Heart Disease: ICD-10: I25;<br>Myocardial infarction: ICD-10: I21, I22<br><br>Procedure (DAD & SDS):<br>Coronary Artery Bypass Grafting:<br>CCI procedure codes: 1IJ76<br>CCP procedure codes: 481<br><br>Percutaneous Coronary Intervention:<br>CCI procedure codes: 1IJ50, 1IJ54, 1IJ57GQ<br>CCP procedure codes: 4802, 4803 |

CONFIDENTIAL

| Variable | Definition |
|---|---|
| | Atrial fibrillation:[6]<br>Individuals with 1 hospitalization or 4 MD visits within a year in the past 5 years with the following codes:<br>ICD-9: 427.31, 427.32<br>ICD-10: I48<br>OHIP dxcode: 427 |
| Chronic respiratory disease | Asthma:[7]<br>An ICES-specific asthma database was used to identify patients with asthma, based on 2 or more ambulatory care visits and/or 1 or more hospitalizations. This variable was included a priori as hypothesized to be directly related to COVID-19 infection risk, as a result of its relationship to severe COVID-19 outcomes.[4]<br><br>OHIP<br>OHIP diagnostic code: 493<br><br>DAD<br>ICD-9 diagnostic code: 493<br>ICD-10 diagnostic codes: J45, J46<br><br>Chronic obstructive pulmonary disease (COPD):[8]<br>An ICES-specific COPD database was used to identify patients with COPD, based on 1 or more ambulatory care visits and/or 1 or more hospitalizations. The algorithm to identify COPD patients was only validated in those aged 35 years or older. This variable was included a priori as hypothesized to be directly related to COVID-19 infection risk.[4]<br><br>OHIP<br>OHIP diagnostic codes: 491, 492, 496<br><br>DAD<br>ICD-9 diagnostic codes: 491, 492, 496<br>ICD-10 diagnostic codes: J41, J42, J43, J44 |
| Hypertension | An ICES-specific hypertension database was used to identify patients with hypertension, based on 1 or more DAD diagnoses or 2 or more OHIP diagnoses in a two-year period; or 1 OHIP diagnosis followed by an OHIP/DAD diagnosis within two years.[9] This variable was included *a priori* as hypothesized to be directly related to COVID-19 infection risk.[4]<br><br>DAD, SDS: ICD-9: 401, 402, 403, 404, 405; ICD-10: I10, I11, I12, I13, I15<br><br>OHIP diagnostic codes: 401, 402, 403, 404, or 405 |
| Diabetes | An ICES-specific diabetes database was used to identify patients with diabetes, based on 2 OHIP diagnostic codes or 1 OHIP service code or 1 DAD admission within 2 years.[10] This variable was included *a priori* as hypothesized to be directly related to COVID-19 infection risk.[4]<br><br>DAD, SDS: ICD-9: 250; ICD-10: E10, E11, E13, E14<br>OHIP: 250<br>OHIP service codes: Q040, K029, K030, K045, K046 |

CONFIDENTIAL

| Variable | Definition |
|---|---|
| Immunocompromised (HIV, transplant, immunosuppressive therapy) | We included immunosuppressive conditions *a priori* as hypothesized to be directly related to COVID-19 infection risk.[4]<br><br><u>HIV:</u>[11]<br>An ICES-specific HIV database was used to identify patients with HIV, based on 3 physician claims in 3 years with OHIP diagnostic codes: 042, 043 or 044<br><br><u>Solid organ transplant recipients:</u><br>CORRLINK is an ICES-specific database which links CORR (Canadian Organ Replacement Register) and DAD data. This database only includes patients that have received an organ transplant and does not include dialysis patients.<br>• For transplants before December 31, 2019: individuals are a transplant recipient if they have a treatment code of 171, where the treatment was before the index date<br>• For transplants on/after January 1, 2020: Identify ICD-10 codes, CCI procedure codes, and OHIP feecodes from DAD, NACRS, and OHIP (codes available upon request)<br><br><u>Allogenic/autologous bone marrow transplant recipients:</u><br>We identified those who had a history of allogenic bone marrow transplant before the index date using the following combination of diagnostic codes:<br>DAD:<br>• CCP procedure codes = 53.0<br>• CCI procedure codes = 1WY19, 1LZ19HHU7, 1LZ19HHU8<br>OHIP:<br>• Feecode = Z426<br><br><u>Other immune disorders</u>:<br>Individuals were identified as having disorders of the immune system based on health care encounters recorded in DAD, SDS, NACRS, and OHIP in the 2-years prior to index using Expanded Diagnostic Clusters from the Johns Hopkins ACGⓇ System Version 10.[12]<br><br><u>Any hospitalization (any diagnosis field) with the following codes</u>:<br>• Sickle-cell disease (ICD-10 D57.0 – D57.2; D57.8 OR ICD-9 282.6);<br>• Other immune system disorders (ICD-9 273.2, 279.0, 279.1, 279.2, 279.3, 279.8, 279.9, 289.8; ICD-10 D80, D81, D82, D83, D84, D89; OHIP dxcode 279)<br>• Immunosuppressive therapy (>30 days (total days supplied) of oral corticosteroid in the 6 months before index date; receipt of other immunocompromising drug, including antineoplastics, in the 6 months before index date)<br><br><u>Active cancer:</u><br>• Any of the following treatments in the past 6 months: cancer surgery (codes available upon request), radiation (if the ICD-10 code listed was Z510 in NACRS), chemotherapy (if the ICD-10 code listed was Z511 or Z512 and any evidence of cancer diagnosis in the Ontario Cancer Registry (OCR) prior to the last treatment date) |

CONFIDENTIAL

| Variable | Definition |
|---|---|
| | • If not any of the above, individuals still classified as having cancer if they had a cancer diagnosis in OCR in the year before the index date |
| Autoimmune disease | Individuals considered to have autoimmune disease if they had any of the following:<br>• Rheumatoid arthritis (identified in the Ontario Rheumatoid Arthritis Database)[13,14]<br>• Inflammatory bowel disease (identified in the Ontario Crohn's and Colitis Cohort)[15,16]<br>• Psoriasis/psoriatic arthritis[17]<br> o Psoriasis: 1 hospitalization or 3 physician billings prior to index date. DAD: ICD-9: 696.1, 696.8; ICD-10: L40.0, L40.1, L40.2, L40.3, L40.4, L40.8, L40.9. OHIP: dxcode = 696<br> o Psoriatic arthritis: 1 hospitalization or: (3 physician billings for psoriatic arthritis + 1 billing for psoriasis [696]). DAD: ICD-9: 696.0; ICD-10: L40.5, M07.0, M07.1, M07.2, M07.3, M09.0. OHIP: dxcode = 721 (at least one of these billings must be billed by a rheumatologist).<br>• Multiple sclerosis[18]<br> o Individuals with one hospitalization or 5 physician billings over 2 years. DAD: ICD-9: 340; ICD-10: G35. OHIP: dxcode = 340. |
| Chronic kidney disease (CKD) | This variable was included *a priori* as hypothesized to be directly related to COVID-19 infection risk.[4] We defined this variable as having a CKD diagnosis code in DAD, NACRS, OHIP in the past 5 years, <u>or:</u> at least 1 dialysis code in each of the 3 months prior to index.[19]<br><br>OHIP: 403, 585<br><br>ICD-10: E102, E112, E132, E142, I12, I13, N08, N18, N19 |
| | Patients who were on chronic dialysis in the year before index date, identified as those with at least 2 of any of the following codes in OHIP, DAD, or SDS separated by at least 90 days, but less than 150 days:[20]<br><br>OHIP service codes: R849, G323, G325, G326, G860, G862, G865 G863, G866, G330, G331, G332, G333, G861, G082, G083, G085, G090, G091, G092, G093, G094, G095, G096, G294, G295, G864, H540, H740<br><br>DAD, SDS:<br>CCI procedure codes: 5195, 6698<br>CCP procedure code: 1PZ21 |
| Advanced liver disease (Cirrhosis or Decompensated Cirrhosis) | We included advanced liver disease *a priori* as hypothesized to be directly related to COVID-19 infection risk.[4]<br><br>Defined using the Cirrhosis Algorithm 9 [from [21]]:<br>Two or more physician visits (diagnosis code 571), or<br>one or more hospital diagnosis of cirrhosis, using the following diagnostic codes:<br>ICD-9 : 456.1, 571.2, 571.5 |

CONFIDENTIAL

| Variable | Definition |
|---|---|
| | ICD-10: I85.9, I98.2, K70.3,K71.7, K74.6<br><br>Defined using the Decompensated Cirrhosis Algorithm 5 (from [21]): One or more physician visits with diagnosis code 571 and (one or more hospital diagnosis or one or more procedure), using the following diagnostic codes:<br><br>ICD-9: 456.0, 456.2, 572.2, 572.3, 572.4, 782.4, 789.5l; ICD-10: I85.0, I86.4, I98.20, I98.3, K721, K729, K76.6, K76.7, R17, R18<br><br>CCI: 1.NA.13.BA-FA, 1.NA.13.BA-X7, 1.NA.13.BA-BD, 1.KQ.76GP-NR, 1.OT.52.HA<br><br>CCP: 1006, 6691<br><br>OHIP: J057, Z591 |
| Dementia | <u>Dementia (ICES cohort) definition:</u><br>1 hospitalization for dementia and/or 3 ambulatory visits for dementia, each separated by at least 30 days, within 2 years and/or 1 prescription from ODB.[22] his variable was included *a priori* as hypothesized to be directly related to COVID-19 infection risk,[4] as well as a marker for healthcare access, mobility, and household-level exposures.[23,24]<br><br>OHIP: 290, 331<br><br>DAD, SDS:<br>ICD-9: 0461, 290.0, 290.1, 290.2, 290.3, 290.4, 294, 331.0, 331.1, 331.5<br>ICD-10: F00, F01, F02, F03, G30<br><br>ODB:<br>1 prescription for a cholinesterase inhibitor |
| Frailty | <u>Frailty:</u><br>Individuals were identified as having medical conditions associated with frailty based on health care encounters recorded in DAD, SDS, NACRS, and OHIP in the 2-years prior to index using Special Population Markers from the Johns Hopkins ACGⓇ System Version 10.[12] |
| History of transient ischemic attack or acute ischemic stroke | This variable was included *a priori* as hypothesized to be directly related to COVID-19 infection risk.[4]<br><br><u>Transient Ischemic Attack:</u><br>DAD and NACRS were used to identify patients with a history of a transient ischemic attack, based on at least 1 hospitalization or ED visit with a diagnosis coded with one of the following codes:<br><br>ICD-9: 435, 3623<br>ICD-10: G450, G451, G452, G453, G458, G459, H340<br><br><u>Acute Ischemic Stroke:</u><br>DAD was used to identify patients with a history of acute ischemic stroke, based on at least 1 hospitalization with a main diagnosis coded with one of the following codes: |

CONFIDENTIAL

| Variable | Definition |
|---|---|
| | ICD-9: 434, 436; ICD-10: I63, I64, H34.1 |
| Influenza vaccine received, 2019-2020 season or 2020-2021 season | An OHIP billing with any of the following fee codes from October 1, 2019 to September 30, 2020, or October 1, 2020 up to 14 days before the index date: G590, G591, G592, Q130, Q590, Q690, Q691;<br><br>or, an ODB billing with any of the following Drug Identification Numbers (DINs)/Product Identification Number (PINs) from October 1, 2019 up to September 30, 2020: 02420643, 02420783, 02432730, 02473283, or October 1, 2020 up to 14 days before the index date: 02420643, 02420783, 02432730, 02445646, 02494248, 09857645, 09857646 |
| Public health unit region | Taken from Public Health Unit (PHU) information using postal code of residence as recorded in the Registered Persons Database and Statistics Canada Postal Code Conversion File Plus (version 7B). Regions were defined as follows:<br><u>Central East</u>: PHU 35 (Haliburton, Kawartha, Pine Ridge District Health Unit), 55 (Peterborough County—City Health Unit), 60 (Simcoe Muskoka District Health Unit)<br><u>Central West</u>: PHU 27 (Brant County Health Unit), 34 (Haldimand-Norfolk Health Unit), 36 (Halton Regional Health Unit), 37 (City of Hamilton Health Unit), 46 (Niagara Regional Area Health Unit), 65 (Waterloo Health Unit), 66 (Wellington-Dufferin-Guelph Health Unit)<br><u>Durham</u>: PHU 30 (Durham Regional Health Unit)<br><u>Eastern</u>: PHU 38 (Hastings and Prince Edward Counties Health Unit), 41 (Kingston, Frontenac and Lennox and Addington Health Unit), 43 (Leeds, Grenville and Lanark District Health Unit), 57 (Renfrew County and District Health Unit), 58 (The Eastern Ontario Health Unit)<br><u>North</u>: PHU 26 (The District of Algoma Health Unit), 47 (North Bay Parry Sound District Health Unit), 49 (Northwestern Health Unit), 56 (Porcupine Health Unit), 61 (Sudbury and District Health Unit), 62 (Thunder Bay District Health Unit), 63 (Timiskaming Health Unit)<br><u>Ottawa</u>: PHU 51 (City of Ottawa Health Unit)<br><u>Peel</u>: PHU 53 (Peel Regional Health Unit)<br><u>South West</u>: PHU 31 (Elgin-St. Thomas), 33 (Grey Bruce Health Unit), 39 (Huron County Health Unit), 40 (Chatham-Kent Health Unit), 42 (Lambton Health Unit), 44 (Middlesex-London Health Unit), 52 (Oxford), 54 (Perth District Health Unit), 68 (Windsor-Essex County Health Unit), 75 (Southwestern Health Unit)<br><u>Toronto</u>: PHU 95 (City of Toronto Health Unit)<br><u>York</u>: PHU 70 (York Regional Health Unit) |
| Dissemination area (DA) | A dissemination area (DA) is the smallest standard geographic area for which all census data are disseminated. A DA generally comprises approximately 400-700 people, but in densely populated cities may contain several thousand people. DAs cover all the territory of Canada.[25]<br>We assigned subjects to a DA using postal code, as recorded in the Registered Persons Database. |
| Household income quintile | Calculated at the DA level using 2016 Census data by multiplying the median income (before-tax) by the number of households and dividing by the sum of single-person equivalent to obtain income per single person equivalent.[26] For DAs where median income was unavailable, |

CONFIDENTIAL

| Variable | Definition |
|---|---|
| | neighbouring DAs were used to estimate income per single person equivalent. DA-based income quintiles were constructed separately for each census metropolitan area or census agglomeration (one or more adjacent municipalities integrated via commuting flows). DAs within each such area were ranked from the lowest average income per single-person equivalent to the highest, and DAs were assigned to five groups, such that each group contained approximately one-fifth the total in-scope population of each area. |
| Persons per dwelling quintile | Average number of persons in private households, calculated at the DA level using the 2016 Census data.[27] DAs across the province were ranked by average number of persons per household into 5 categories (quintiles), such that each group contained approximately one-fifth of the DAs. |
| Essential worker quintile | Calculated at the DA level, using 2016 Census data.[28] For each DA, we calculated the number of individuals ≥15 years old that were working in one of the following Census-defined work categories: Sales and service occupations; trades, transport and equipment operators and related occupations; natural resources, agriculture and related production occupations; and occupations in manufacturing and utilities.<br><br>DAs across the province were then ranked by these percentages into quintiles, with the lowest 1/5 of DAs comprising the first quintile, and so on. |
| Visible minority quintile | Calculated at the DA level, using 2016 Census data.[28] An individual was marked as “self-identify as a visible minority” if they reported being one or more of the following (wording from the 2016 Census): “South Asian (e.g., East Indian, Pakistani, Sri Lankan, etc.), Chinese, Black, Filipino, Latin American, Arab, Southeast Asian (e.g., Vietnamese, Cambodian, Laotian, Thai, etc.), West Asian (e.g., Iranian, Afghan, etc.), Korean, Japanese, or Other—specify”.<br><br>DAs across the province were then ranked by these percentages into quintiles, with the lowest 1/5 of DAs comprising the first quintile, and so on. |

CONFIDENTIAL

**eTable 2:** Comparison of unadjusted vaccine effectiveness estimates and VE estimates adjusted for a single covariate to evaluate the impact of potential confounders.

| | **Symptomatic infection** | | | | **Severe outcomes** | | | |
|---|---|---|---|---|---|---|---|---|
| | ≥14 days after Dose 1[a] | | ≥7 days after Dose 2 | | ≥14 days after Dose 1[a] | | ≥7 days after Dose 2 | |
| | VE (95% CI) | % change in OR[b] | VE (95% CI) | % change in OR** | VE (95% CI) | % change in OR[b] | VE (95% CI) | % change in OR** |
| **Unadjusted** | **60 (57, 63)** | **-** | **93 (91, 94)** | **-** | **25 (3, 43)** | **-** | **97 (80, 100)** | **-** |
| Adjusted for: | | | | | | | | |
| Age group | 58 (55, 61) | 5.0 | 92 (90, 94) | 3.9 | 60 (48, 70) | -47 | 98 (88, 100) | -39 |
| Sex | 59 (55, 62) | 4.2 | 92 (90, 94) | 5.9 | 18 (-7, 37) | 9.9 | 97 (77, 100) | 14 |
| Public Health Unit Region | 58 (54, 61) | 6.7 | 93 (91, 95) | -4.9 | 23 (0, 41) | 3.5 | 97 (81, 100) | -4.3 |
| Biweekly period of test | 72 (69, 74) | -29 | 94 (92, 95) | -12 | 33 (12, 48) | -9.6 | 97 (79, 100) | 2.3 |
| Number of tests in previous 3 months | 56 (53, 59) | 10 | 90 (87, 93) | 32 | 19 (-5, 38) | 7.8 | 97 (76, 100) | 21 |
| Any comorbidity | 60 (56, 63) | 1.7 | 93 (90, 94) | 0.9 | 34 (15, 50) | -12 | 97 (81, 100) | -7.3 |
| Receipt of 2019-2020 and/or 2020-2021 influenza vaccination | 58 (55, 61) | 5.4 | 93 (90, 94) | 1.6 | 31 (10, 47) | -7.1 | 97 (80, 100) | -2.3 |
| Household income quintile | 60 (57, 63) | 0.3 | 93 (91, 94) | 0.1 | 24 (2, 42) | 0.7 | 97 (80, 100) | 0.4 |
| Essential workers quintile | 60 (57, 63) | -0.6 | 93 (90, 94) | 1.4 | 26 (4, 43) | -1.1 | 97 (79, 100) | 1.8 |
| Persons per dwelling quintile | 58 (54, 61) | 6.2 | 93 (90, 94) | 0.5 | 24 (1, 41) | 2.4 | 97 (80, 100) | 0.0 |
| Self-identified visible minorities quintile | 55 (52, 59) | 12 | 93 (91, 95) | -2.1 | 18 (-7, 37) | 10 | 97 (80, 100) | -2.0 |
| **Adjusted for all covariates** | **60 (57, 64)** | **-0.6** | **91 (89, 93)** | **20** | **70 (60, 77)** | **-59** | **98 (88, 100)** | **-43** |

[a]Among individuals who received only one dose by the index date
[b]% Change in OR = (adjusted OR/crude OR - 1)*100

CONFIDENTIAL

**eFigure 1:** Distribution of mRNA vaccine product (BNT162b2, mRNA-1273) among vaccinated individuals included in the study cohort (Panel A) and all adults aged ≥16 years in Ontario, Canada (Panels B, C) between 14 December 2020 and 19 April 2021.

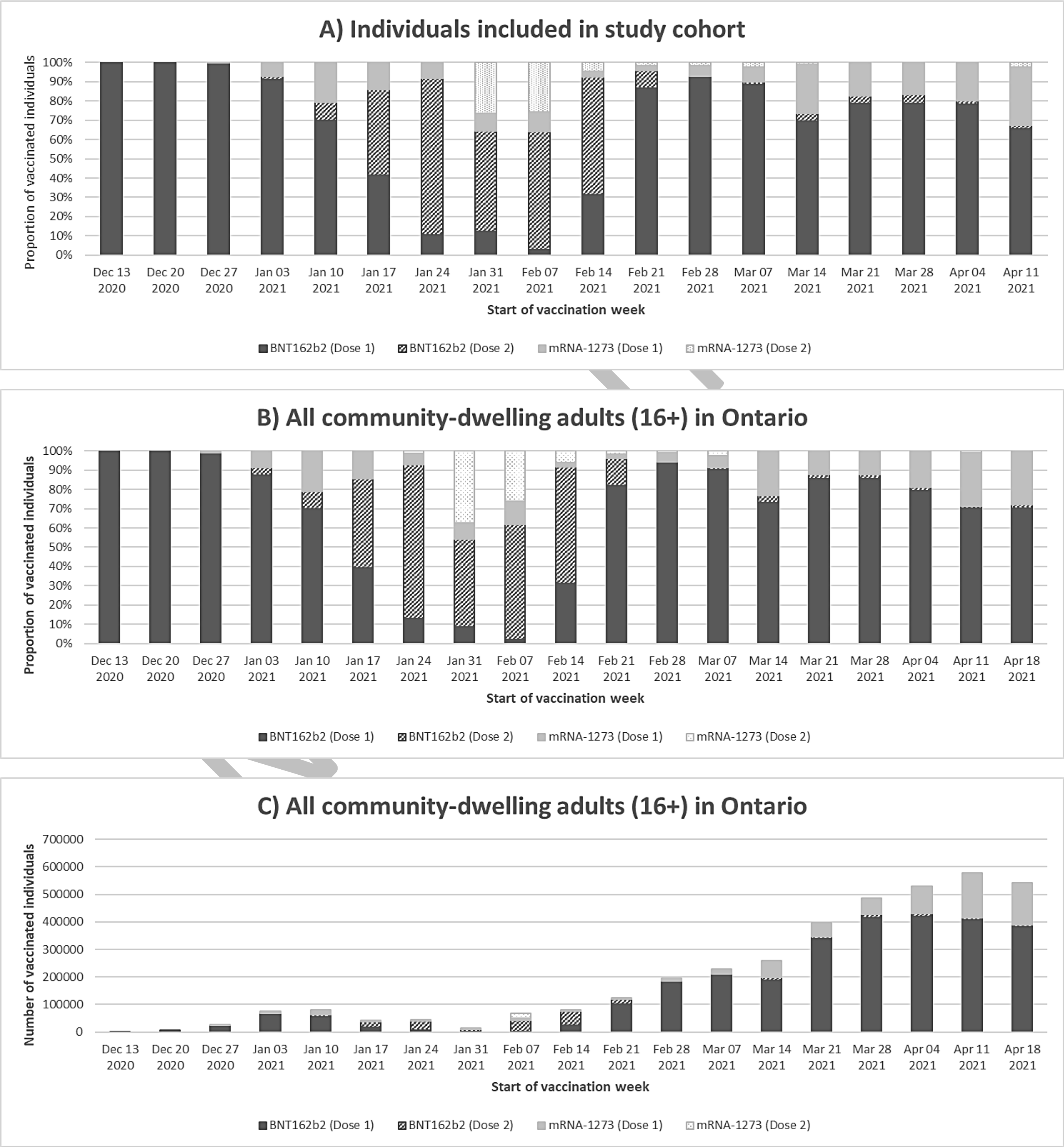

**eTable 3:** Characteristics of individuals with symptoms consistent with COVID-19 recorded in the Ontario Laboratories Information System (OLIS) at the time of testing for SARS-CoV-2, individuals recorded in OLIS as asymptomatic, and individuals who did not have symptoms consistent with COVID-19 or had no symptom information recorded in OLIS between 14 December 2020 and 19 April 2021 in Ontario, Canada.

| Characteristic | (A) COVID-19 symptoms, n (%)[a] (N=324,033) | (B) Asymptomatic, n (%)[a] (N=519,885) | (C) COVID-19 symptoms and asymptomatic combined, n (%)[a] (N=843,918) | (D) Recorded symptoms not consistent with COVID-19 or no symptom information recorded, n (%)[a] (N=1,293,768) | Standardized difference[b] (C vs. D) |
|---|---|---|---|---|---|
| Received ≥1 dose of COVID-19 vaccine | 21,272 (6.6) | 83,890 (16.1) | 105,162 (12.5) | 93,639 (7.2) | 0.18 |
| Tested positive for SARS-CoV-2 | 53,270 (16.4) | 22,740 (4.4) | 76,010 (9.0) | 136,653 (10.6) | 0.05 |
| Age (years), mean (standard deviation) | 43.0 (17.7) | 47.0 (18.9) | 45.5 (18.5) | 45.8 (18.6) | 0.01 |
| Age group (years) | | | | | |
| 16–29 | 87,413 (27.0) | 113,375 (21.8) | 200,788 (23.8) | 303,837 (23.5) | 0.01 |
| 30–39 | 69,350 (21.4) | 87,079 (16.7) | 156,429 (18.5) | 247,433 (19.1) | 0.02 |
| 40–49 | 55,867 (17.2) | 88,466 (17.0) | 144,333 (17.1) | 211,122 (16.3) | 0.02 |
| 50–59 | 50,334 (15.5) | 97,557 (18.8) | 147,891 (17.5) | 215,614 (16.7) | 0.02 |
| 60–69 | 32,621 (10.1) | 69,504 (13.4) | 102,125 (12.1) | 159,036 (12.3) | 0.01 |
| 70–79 | 17,314 (5.3) | 32,322 (6.2) | 49,636 (5.9) | 94,374 (7.3) | 0.06 |
| ≥80 | 11,134 (3.4) | 31,582 (6.1) | 42,716 (5.1) | 62,352 (4.8) | 0.01 |
| Male sex | 138,494 (42.7) | 208,819 (40.2) | 347,313 (41.2) | 585,559 (45.3) | 0.08 |
| Public health unit region[c] | | | | | |
| Central East | 31,818 (9.8) | 47,792 (9.2) | 79,610 (9.4) | 63,357 (4.9) | 0.18 |
| Central West | 56,741 (17.5) | 86,609 (16.7) | 143,350 (17.0) | 271,921 (21.0) | 0.10 |
| Durham | 10,016 (3.1) | 19,991 (3.8) | 30,007 (3.6) | 70,212 (5.4) | 0.09 |
| Eastern | 15,836 (4.9) | 24,630 (4.7) | 40,466 (4.8) | 90,671 (7.0) | 0.09 |
| North | 33,074 (10.2) | 46,562 (9.0) | 79,636 (9.4) | 48,150 (3.7) | 0.23 |
| Ottawa | 3,561 (1.1) | 15,029 (2.9) | 18,590 (2.2) | 112,651 (8.7) | 0.29 |
| Peel | 46,496 (14.3) | 61,058 (11.7) | 107,554 (12.7) | 107,572 (8.3) | 0.14 |
| South West | 46,878 (14.5) | 73,947 (14.2) | 120,825 (14.3) | 126,449 (9.8) | 0.14 |
| Toronto | 57,998 (17.9) | 98,517 (18.9) | 156,515 (18.5) | 282,852 (21.9) | 0.08 |
| York | 20,273 (6.3) | 43,618 (8.4) | 63,891 (7.6) | 115,422 (8.9) | 0.05 |
| Biweekly period of test | | | | | |
| 14 Dec 2020 to 27 Dec 2020 | 31,623 (9.8) | 51,533 (9.9) | 83,156 (9.9) | 172,097 (13.3) | 0.11 |
| 28 Dec 2020 to 10 Jan 2021 | 33,825 (10.4) | 50,050 (9.6) | 83,875 (9.9) | 164,818 (12.7) | 0.09 |

CONFIDENTIAL

| Characteristic | (A) COVID-19 symptoms, n (%)[a] (N=324,033) | (B) Asymptomatic, n (%)[a] (N=519,885) | (C) COVID-19 symptoms and asymptomatic combined, n (%)[a] (N=843,918) | (D) Recorded symptoms not consistent with COVID-19 or no symptom information recorded, n (%)[a] (N=1,293,768) | Standardized difference[b] (C vs. D) |
|---|---|---|---|---|---|
| 11 Jan 2021 to 24 Jan 2021 | 31,600 (9.8) | 48,456 (9.3) | 80,056 (9.5) | 147,371 (11.4) | 0.06 |
| 25 Jan 2021 to 7 Feb 2021 | 27,852 (8.6) | 46,513 (8.9) | 74,365 (8.8) | 118,455 (9.2) | 0.01 |
| 8 Feb 2021 to 21 Feb 2021 | 28,302 (8.7) | 54,532 (10.5) | 82,834 (9.8) | 103,879 (8.0) | 0.06 |
| 22 Feb 2021 to 7 Mar 2021 | 34,367 (10.6) | 61,588 (11.8) | 95,955 (11.4) | 114,039 (8.8) | 0.08 |
| 8 Mar 2021 to 21 Mar 2021 | 37,969 (11.7) | 61,008 (11.7) | 98,977 (11.7) | 124,445 (9.6) | 0.07 |
| 22 Mar 2021 to 4 Apr 2021 | 44,218 (13.6) | 67,522 (13.0) | 111,740 (13.2) | 146,394 (11.3) | 0.06 |
| 5 Apr 2021 to 19 Apr 2021 | 54,277 (16.8) | 78,683 (15.1) | 132,960 (15.8) | 202,270 (15.6) | 0.00 |
| Number of tests in previous 3 months | | | | | |
| 0 | 233,499 (72.1) | 312,611 (60.1) | 546,110 (64.7) | 969,250 (74.9) | 0.22 |
| 1 | 61,978 (19.1) | 77,354 (14.9) | 139,332 (16.5) | 222,312 (17.2) | 0.02 |
| ≥2 | 28,556 (8.8) | 129,920 (25.0) | 158,476 (18.8) | 102,206 (7.9) | 0.32 |
| Any comorbidity[d] | 151,186 (46.7) | 250,466 (48.2) | 401,652 (47.6) | 619,430 (47.9) | 0.01 |
| Receipt of 2019-2020 and/or 2020-2021 influenza vaccination | 103,146 (31.8) | 174,271 (33.5) | 277,417 (32.9) | 437,059 (33.8) | 0.02 |
| Household income quintile[c, e] | | | | | |
| 1 (lowest) | 59,822 (18.5) | 103,771 (20.0) | 163,593 (19.4) | 251,573 (19.4) | 0.00 |
| 2 | 62,624 (19.3) | 102,050 (19.6) | 164,674 (19.5) | 250,720 (19.4) | 0.00 |
| 3 | 64,105 (19.8) | 103,053 (19.8) | 167,158 (19.8) | 261,983 (20.2) | 0.01 |
| 4 | 66,822 (20.6) | 104,840 (20.2) | 171,662 (20.3) | 261,363 (20.2) | 0.00 |
| 5 (highest) | 69,133 (21.3) | 103,711 (19.9) | 172,844 (20.5) | 263,195 (20.3) | 0.00 |
| Essential workers quintile[c, f] | | | | | |
| 1 (0%–32.5%) | 57,104 (17.6) | 86,085 (16.6) | 143,189 (17.0) | 294,015 (22.7) | 0.14 |
| 2 (32.5%–42.3%) | 71,265 (22.0) | 114,051 (21.9) | 185,316 (22.0) | 290,191 (22.4) | 0.01 |
| 3 (42.3%–49.8%) | 67,214 (20.7) | 109,473 (21.1) | 176,687 (20.9) | 253,246 (19.6) | 0.03 |
| 4 (50.0%–57.5%) | 64,425 (19.9) | 106,133 (20.4) | 170,558 (20.2) | 240,766 (18.6) | 0.04 |
| 5 (57.5%–100%) | 61,586 (19.0) | 99,835 (19.2) | 161,421 (19.1) | 209,013 (16.2) | 0.08 |
| Persons per dwelling quintile[c, g] | | | | | |
| 1 (0–2.1) | 57,633 (17.8) | 89,760 (17.3) | 147,393 (17.5) | 246,369 (19.0) | 0.04 |
| 2 (2.2–2.4) | 58,967 (18.2) | 92,229 (17.7) | 151,196 (17.9) | 217,568 (16.8) | 0.03 |
| 3 (2.5–2.6) | 42,862 (13.2) | 69,632 (13.4) | 112,494 (13.3) | 180,832 (14.0) | 0.02 |
| 4 (2.7–3.0) | 76,741 (23.7) | 125,991 (24.2) | 202,732 (24.0) | 307,590 (23.8) | 0.01 |

CONFIDENTIAL

| Characteristic | (A) COVID-19 symptoms, n (%)[a] (N=324,033) | (B) Asymptomatic, n (%)[a] (N=519,885) | (C) COVID-19 symptoms and asymptomatic combined, n (%)[a] (N=843,918) | (D) Recorded symptoms not consistent with COVID-19 or no symptom information recorded, n (%)[a] (N=1,293,768) | Standardized difference[b] (C vs. D) |
|---|---|---|---|---|---|
| 5 (3.1–5.7) | 85,292 (26.3) | 137,732 (26.5) | 223,024 (26.4) | 334,154 (25.8) | 0.01 |
| Self-identified visible minority quintile[c, h] | | | | | |
| 1 (0.0%–2.2%) | 56,356 (17.4) | 92,897 (17.9) | 149,253 (17.7) | 180,221 (13.9) | 0.10 |
| 2 (2.2%–7.5%) | 60,876 (18.8) | 92,318 (17.8) | 153,194 (18.2) | 210,399 (16.3) | 0.05 |
| 3 (7.5%–18.7%) | 58,345 (18.0) | 91,061 (17.5) | 149,406 (17.7) | 244,332 (18.9) | 0.03 |
| 4 (18.7%–43.5%) | 64,409 (19.9) | 102,903 (19.8) | 167,312 (19.8) | 304,299 (23.5) | 0.09 |
| 5 (43.5%–100%) | 81,612 (25.2) | 136,413 (26.2) | 218,025 (25.8) | 348,009 (26.9) | 0.02 |

[a]Proportion reported, unless stated otherwise.
[b]Standardized differences of >0.10 are considered clinically relevant.
[c]The sum of counts does not equal the column total because of individuals with missing information (<1.0%) for this characteristic.
[d]Comorbidities include chronic respiratory diseases, chronic heart diseases, hypertension, diabetes, immunocompromising conditions due to underlying diseases or therapy, autoimmune diseases, chronic kidney disease, advanced liver disease, dementia/frailty and history of stroke or transient ischemic attack.
[e]Household income quintile has variable cut-off values in each city/Census area to account for cost of living. A dissemination area (DA) being in quintile 1 means it is among the lowest 20% of DAs in its city by income.
[f]Percentage of people in the area working in the following occupations: sales and service occupations; trades, transport and equipment operators and related occupations; natural resources, agriculture, and related production occupations; and occupations in manufacturing and utilities. Census counts for people are randomly rounded up or down to the nearest number divisible by 5, which causes some minor imprecision.
[g]Range of persons per dwelling.
[h]Percentage of people in the area who self-identified as a visible minority. Census counts for people are randomly rounded up or down to the nearest number divisible by 5, which causes some minor imprecision.

CONFIDENTIAL

**eTable 4:** Characteristics of symptomatic individuals tested for SARS-CoV-2 according to vaccine product and vaccination status between 14 December 2020 and 19 April 2021 in Ontario, Canada.

| Characteristic | BNT162b2 | | mRNA-1273 | |
|---|---|---|---|---|
| | Received only 1 dose, n (%)[a] (N= 14,156) | Received 2 doses, n (%)[a] (N= 4,176) | Received only 1 dose, n (%)[a] (N= 2,222) | Received 2 doses, n (%)[a] (N= 718) |
| Interval between receipt of only dose 1 and index date, or interval between dose 2 and index date (days), median (IQR) | 16 (8–27) | 30 (10–51) | 12 (6–21) | 26 (7–44) |
| Interval between 2 doses (days), median (IQR) | - | 25 (21–35) | - | 28 (28–28) |
| Age (years), mean (standard deviation) | 51.8 (20.4) | 45.6 (18.6) | 57.7 (21.6) | 68.4 (23.1) |
| Age group (years) | | | | |
| 16–29 | 2,292 (16.2) | 854 (20.5) | 253 (11.4) | 58 (8.1) |
| 30–39 | 2,592 (18.3) | 1,035 (24.8) | 328 (14.8) | 56 (7.8) |
| 40–49 | 2,131 (15.1) | 799 (19.1) | 293 (13.2) | 64 (8.9) |
| 50–59 | 2,010 (14.2) | 719 (17.2) | 314 (14.1) | 69 (9.6) |
| 60–69 | 1,653 (11.7) | 304 (7.3) | 242 (10.9) | 62 (8.6) |
| 70–79 | 1,774 (12.5) | 99 (2.4) | 318 (14.3) | 81 (11.3) |
| ≥80 | 1,704 (12.0) | 366 (8.8) | 474 (21.3) | 328 (45.7) |
| Male sex | 4,152 (29.3) | 882 (21.1) | 774 (34.8) | 205 (28.6) |
| Public health unit region[b] | | | | |
| Central East | 1,381 (9.8) | 519 (12.4) | 53 (2.4) | 16 (2.2) |
| Central West | 2,495 (17.6) | 1,044 (25.0) | 220 (9.9) | 94 (13.1) |
| Durham | 369 (2.6) | 112 (2.7) | 33 (1.5) | 8 (1.1) |
| Eastern | 834 (5.9) | 142 (3.4) | 72 (3.2) | 39 (5.4) |
| North | 1,366 (9.6) | 112 (2.7) | 735 (33.1) | 38 (5.3) |
| Ottawa | 258 (1.8) | 147 (3.5) | 22 (1.0) | 19 (2.6) |
| Peel | 1,499 (10.6) | 500 (12.0) | 272 (12.2) | 124 (17.3) |
| South West | 2,840 (20.1) | 599 (14.3) | 302 (13.6) | 144 (20.1) |
| Toronto | 2,170 (15.3) | 756 (18.1) | 357 (16.1) | 179 (24.9) |
| York | 892 (6.3) | 231 (5.5) | 151 (6.8) | 49 (6.8) |
| Biweekly period of test | | | | |
| 14 Dec 2020 to 27 Dec 2020 | 13 (0.1) | 0 (0.0) | 0 (0.0) | 0 (0.0) |
| 28 Dec 2020 to 10 Jan 2021 | 320 (2.3) | 8 (0.2) | 7 (0.3) | 0 (0.0) |
| 11 Jan 2021 to 24 Jan 2021 | 782 (5.5) | 151 (3.6) | 135 (6.1) | 0 (0.0) |
| 25 Jan 2021 to 7 Feb 2021 | 657 (4.6) | 394 (9.4) | 137 (6.2) | 16 (2.2) |
| 8 Feb 2021 to 21 Feb 2021 | 313 (2.2) | 523 (12.5) | 77 (3.5) | 118 (16.4) |
| 22 Feb 2021 to 7 Mar 2021 | 597 (4.2) | 681 (16.3) | 86 (3.9) | 127 (17.7) |

CONFIDENTIAL

| Characteristic | BNT162b2 | | mRNA-1273 | |
|---|---|---|---|---|
| | Received only 1 dose, n (%)[a] (N= 14,156) | Received 2 doses, n (%)[a] (N= 4,176) | Received only 1 dose, n (%)[a] (N= 2,222) | Received 2 doses, n (%)[a] (N= 718) |
| 8 Mar 2021 to 21 Mar 2021 | 1,820 (12.9) | 665 (15.9) | 179 (8.1) | 126 (17.5) |
| 22 Mar 2021 to 4 Apr 2021 | 3,388 (23.9) | 776 (18.6) | 513 (23.1) | 137 (19.1) |
| 5 Apr 2021 to 19 Apr 2021 | 6,266 (44.3) | 978 (23.4) | 1,088 (49.0) | 194 (27.0) |
| Number of tests in previous 3 months | | | | |
| 0 | 8,546 (60.4) | 1,489 (35.7) | 1,331 (59.9) | 222 (30.9) |
| 1 | 2,859 (20.2) | 902 (21.6) | 435 (19.6) | 142 (19.8) |
| ≥2 | 2,751 (19.4) | 1,785 (42.7) | 456 (20.5) | 354 (49.3) |
| Any comorbidity[c] | 8,159 (57.6) | 2,002 (47.9) | 1,496 (67.3) | 561 (78.1) |
| Receipt of 2019-2020 and/or 2020-2021 influenza vaccination | 6,878 (48.6) | 1,361 (32.6) | 978 (44.0) | 370 (51.5) |
| Household income quintile[b, d] | | | | |
| 1 (lowest) | 2,416 (17.1) | 667 (16.0) | 532 (23.9) | 135 (18.8) |
| 2 | 2,690 (19.0) | 819 (19.6) | 459 (20.7) | 178 (24.8) |
| 3 | 2,842 (20.1) | 816 (19.5) | 443 (19.9) | 132 (18.4) |
| 4 | 3,037 (21.5) | 928 (22.2) | 401 (18.0) | 147 (20.5) |
| 5 (highest) | 3,112 (22.0) | 930 (22.3) | 380 (17.1) | 118 (16.4) |
| Essential workers quintile[b, e] | | | | |
| 1 (0%–32.5%) | 2,527 (17.9) | 924 (22.1) | 308 (13.9) | 158 (22.0) |
| 2 (32.5%–42.3%) | 3,074 (21.7) | 912 (21.8) | 520 (23.4) | 158 (22.0) |
| 3 (42.3%–49.8%) | 3,001 (21.2) | 868 (20.8) | 451 (20.3) | 148 (20.6) |
| 4 (50.0%–57.5%) | 2,845 (20.1) | 786 (18.8) | 463 (20.8) | 117 (16.3) |
| 5 (57.5%–100%) | 2,612 (18.5) | 658 (15.8) | 462 (20.8) | 127 (17.7) |
| Persons per dwelling quintile[b, f] | | | | |
| 1 (0–2.1) | 2,796 (19.8) | 757 (18.1) | 536 (24.1) | 188 (26.2) |
| 2 (2.2–2.4) | 2,834 (20.0) | 697 (16.7) | 557 (25.1) | 131 (18.2) |
| 3 (2.5–2.6) | 2,058 (14.5) | 577 (13.8) | 290 (13.1) | 95 (13.2) |
| 4 (2.7–3.0) | 3,255 (23.0) | 1,084 (26.0) | 378 (17.0) | 157 (21.9) |
| 5 (3.1–5.7) | 3,108 (22.0) | 1,023 (24.5) | 440 (19.8) | 138 (19.2) |
| Self-identified visible minority quintile[b, g] | | | | |
| 1 (0.0%–2.2%) | 2,768 (19.6) | 594 (14.2) | 670 (30.2) | 101 (14.1) |
| 2 (2.2%–7.5%) | 3,245 (22.9) | 794 (19.0) | 444 (20.0) | 109 (15.2) |
| 3 (7.5%–18.7%) | 2,684 (19.0) | 844 (20.2) | 303 (13.6) | 151 (21.0) |
| 4 (18.7%–43.5%) | 2,507 (17.7) | 950 (22.7) | 344 (15.5) | 173 (24.1) |
| 5 (43.5%–100%) | 2,855 (20.2) | 966 (23.1) | 443 (19.9) | 174 (24.2) |

[a]Proportion reported, unless stated otherwise.
[b]The sum of counts does not equal the column total because of individuals with missing information (<1.0%) for this characteristic.

CONFIDENTIAL

[c]Comorbidities include chronic respiratory diseases, chronic heart diseases, hypertension, diabetes, immunocompromising conditions due to underlying diseases or therapy, autoimmune diseases, chronic kidney disease, advanced liver disease, dementia/frailty and history of stroke or transient ischemic attack.
[d]Household income quintile has variable cut-off values in each city/Census area to account for cost of living. A dissemination area (DA) being in quintile 1 means it is among the lowest 20% of DAs in its city by income.
[e]Percentage of people in the area working in the following occupations: sales and service occupations; trades, transport and equipment operators and related occupations; natural resources, agriculture, and related production occupations; and occupations in manufacturing and utilities. Census counts for people are randomly rounded up or down to the nearest number divisible by 5, which causes some minor imprecision.
[f]Range of persons per dwelling.
[g]Percentage of people in the area who self-identified as a visible minority. Census counts for people are randomly rounded up or down to the nearest number divisible by 5, which causes some minor imprecision.

CONFIDENTIAL

**eTable 5:** Unadjusted and adjusted vaccine effectiveness estimates of COVID-19 mRNA vaccines against symptomatic SARS-CoV-2 infection and severe outcomes (hospitalization or death) by various intervals, between 14 December 2020 and 19 April 2021 in Ontario, Canada.

| | Test-positive cases, No. Vaccinated/Total[a] | Test-negative controls, No. Vaccinated/Total[a] | Unadjusted VE% (95% CI) | Adjusted[b] VE% (95% CI) | Adjusted[c] VE% (95% CI) |
|---|---|---|---|---|---|
| **Symptomatic SARS-CoV-2 infection** | | | | | |
| **Received one dose, time since Dose 1** | | | | | |
| 0–6 days | 520 / 51,740 | 2,674 / 254,215 | 5 (-5, 13) | 3 (-8, 12) | Reference |
| 7–13 days | 772 / 51,992 | 3,282 / 254,823 | -16 (-25, -7) | -16 (-26, -6) | |
| 14–20 days | 283 / 51,503 | 2,794 / 254,335 | 50 (44, 56) | 48 (41, 54) | 50 (43, 57) |
| 21–27 days | 161 / 51,381 | 2,130 / 253,671 | 63 (56, 68) | 60 (53, 66) | 58 (49, 65) |
| 28–34 days | 108 / 51,328 | 1,647 / 253,188 | 68 (61, 74) | 68 (60, 73) | 62 (52, 69) |
| 35–41 days | 72 / 51,292 | 1,063 / 252,604 | 67 (58, 74) | 71 (63, 78) | 67 (57, 74) |
| 42–48 days | 42 / 51,262 | 594 / 252,135 | 65 (53, 75) | 70 (58, 78) | 66 (52, 75) |
| ≥49 days | 19 / 51,239 | 217 / 251,758 | 57 (31, 73) | 64 (42, 78) | 60 (35, 76) |
| **Received two doses, time since Dose 2** | | | | | |
| 0–6 days | 16 / 51,236 | 1,004 / 252,545 | 92 (87, 95) | 88 (81, 93) | 88 (80, 93) |
| ≥7 days | 57 / 51,277 | 3,817 / 255,358 | 93 (91, 94) | 91 (89, 93) | 91 (88, 93) |
| *≥14 days* | Suppressed[d] | Suppressed[d] | 92 (90, 94) | 91 (88, 93) | 91 (87, 93) |
| **COVID-19 hospitalization or death** | | | | | |
| **Received one dose, time since Dose 1** | | | | | |
| 0–6 days | Suppressed[d] | 2,674 / 254,215 | -79 (-142, -32) | 20 (-10, 41) | Reference |
| 7–13 days | Suppressed[d] | 3,282 / 254,823 | -99 (-158, -54) | 26 (2, 43) | |
| 14–20 days | Suppressed[d] | 2,794 / 254,335 | -5 (-54, 28) | 62 (44, 75) | 53 (26, 70) |
| 21–27 days | Suppressed[d] | 2,130 / 253,671 | -2 (-59, 34) | 60 (37, 75) | 48 (13, 69) |
| 28–34 days | Suppressed[d] | 1,647 / 253,188 | 47 (-6, 74) | 76 (52, 88) | 68 (31, 85) |
| ≥35 days | Suppressed[d] | 1,874 / 253,415 | 83 (46, 94) | 91 (73, 97) | 88 (60, 96) |
| **Received two doses, time since Dose 2** | | | | | |
| 0–6 days | Suppressed[d] | 1,004 / 252,545 | 78 (13, 95) | 83 (32, 96) | 78 (-2, 95) |
| ≥7 days | Suppressed[d] | 3,817 / 255,358 | 97 (80, 100) | 98 (88, 100) | 98 (83, 100) |

[a]The denominator represents the total number of test-positive cases or test-negative controls (vaccinated and unvaccinated), whereas the numerator represents only the number of test-positive cases or test-negative controls that are vaccinated.

CONFIDENTIAL

[b]Models were adjusted for age, sex, public health unit region, biweekly period of test, number of SARS-CoV-2 tests in the 3 months prior to 14 December 2020, presence of any comorbidity that increase the risk of severe COVID-19, receipt of influenza vaccination in current or prior influenza season, and neighbourhood-level household income, persons per dwelling, proportion of persons employed as non-health essential workers, and self-identified visible minority quintiles.

[c]Analyses restricted to only individuals who were vaccinated, where individuals vaccinated 0-13 days before index date were considered 'unvaccinated' and served as the reference group.

[d]Counts of vaccinated test-positive cases for each interval are suppressed because they are either small cell counts (≤5, which cannot be disclosed because of privacy and data obligations) or they could be derived using other estimates in this table. Overall, 165/2,479 (6.6%) of test-positive cases were vaccinated.

CONFIDENTIAL

**eTable 6:** Unadjusted and adjusted vaccine effectiveness estimates against symptomatic COVID-19 infection after Dose 1 (for individuals who received only 1 dose) and Dose 2, by various factors, including vaccine product, patient characteristics, epidemic wave, and SARS-CoV-2 lineage between 14 December 2020 and 19 April 2021 in Ontario, Canada.

| | Test-positive cases, No. Vaccinated/Total[a] | Test-negative controls, No. Vaccinated/Total[a] | Unadjusted VE% (95% CI) | Adjusted[b] VE% (95% CI) |
|---|---|---|---|---|
| **Vaccine product** | | | | |
| BNT162b2 | | | | |
| ≥14 days after Dose 1 | 636 / 51,856 | 7,483 / 259,024 | 58 (55, 62) | 59 (55, 62) |
| ≥7 days after Dose 2 | 51 / 51,271 | 3,275 / 254,816 | 92 (90, 94) | 91 (88, 93) |
| *By age group* | | | | |
| ≥70 years | | | | |
| ≥14 days after Dose 1 | Suppressed[c] | Suppressed[c] | -5 (-20, 9) | 40 (29, 50) |
| ≥7 days after Dose 2 | Suppressed[c] | Suppressed[c] | 93 (81, 97) | 93 (82, 98) |
| 40–69 years | | | | |
| ≥14 days after Dose 1 | Suppressed[c] | Suppressed[c] | 66 (61, 71) | 64 (58, 69) |
| ≥7 days after Dose 2 | Suppressed[c] | Suppressed[c] | 90 (86, 93) | 88 (83, 92) |
| 16–39 years | | | | |
| ≥14 days after Dose 1 | Suppressed[c] | Suppressed[c] | 72 (67, 76) | 69 (64, 74) |
| ≥7 days after Dose 2 | Suppressed[c] | Suppressed[c] | 94 (91, 96) | 93 (88, 96) |
| mRNA-1273 | | | | |
| ≥14 days after Dose 1 | 49 / 51,269 | 962 / 252,503 | 75 (67, 81) | 72 (63, 80) |
| ≥7 days after Dose 2 | 6 / 51,226 | 542 / 252,083 | 95 (88, 98) | 94 (86, 97) |
| *By age group* | | | | |
| ≥70 years | | | | |
| ≥14 days after Dose 1 | Suppressed[c] | Suppressed[c] | 41 (15, 59) | 54 (31, 69) |
| ≥7 days after Dose 2 | Suppressed[c] | Suppressed[c] | 94 (80, 98) | 95 (83, 98) |
| 40–69 years | | | | |
| ≥14 days after Dose 1 | Suppressed[c] | Suppressed[c] | 83 (70, 90) | 82 (68, 90) |
| ≥7 days after Dose 2 | Suppressed[c] | Suppressed[c] | 97 (77, 100) | 97 (75, 100) |
| 16–39 years | | | | |
| ≥14 days after Dose 1 | Suppressed[c] | Suppressed[c] | 90 (77, 96) | 89 (73, 95) |
| ≥7 days after Dose 2 | Suppressed[c] | Suppressed[c] | 88 (49, 97) | 86 (44, 97) |
| **Age group** | | | | |
| ≥70 years | | | | |
| ≥14 days after Dose 1 | 285 / 3,308 | 1,977 / 22,258 | 3 (-10, 15) | 40 (29, 49) |
| ≥7 days after Dose 2 | 7 / 3,030 | 690 / 20,971 | 93 (86, 97) | 94 (87, 97) |
| 40–69 years | | | | |
| ≥14 days after Dose 1 | 237 / 23,697 | 3,381 / 110,083 | 68 (64, 72) | 66 (61, 70) |
| ≥7 days after Dose 2 | 31 / 23,491 | 1,574 / 108,276 | 91 (87, 94) | 89 (85, 93) |

CONFIDENTIAL

| | Test-positive cases, No. Vaccinated/Total[a] | Test-negative controls, No. Vaccinated/Total[a] | Unadjusted VE% (95% CI) | Adjusted[b] VE% (95% CI) |
|---|---|---|---|---|
| 16–39 years | | | | |
| ≥14 days after Dose 1 | 163 / 24,900 | 3,087 / 127,645 | 73 (69, 77) | 71 (66, 75) |
| ≥7 days after Dose 2 | 19 / 24,756 | 1,553 / 126,111 | 94 (90, 96) | 93 (88, 95) |
| **Sex** | | | | |
| Male | | | | |
| ≥14 days after Dose 1 | 247 / 25,453 | 2,317 / 109,592 | 55 (48, 60) | 56 (49, 62) |
| ≥7 days after Dose 2 | 9 / 25,215 | 840 / 108,115 | 95 (91, 98) | 94 (89, 97) |
| Female | | | | |
| ≥14 days after Dose 1 | 438 / 26,452 | 6,128 / 150,394 | 60 (56, 64) | 62 (58, 66) |
| ≥7 days after Dose 2 | 48 / 26,062 | 2,977 / 147,243 | 91 (88, 93) | 90 (87, 93) |
| **Presence of any comorbidity** | | | | |
| Yes | | | | |
| ≥14 days after Dose 1 | 457 / 22,349 | 4,788 / 121,864 | 49 (44, 54) | 54 (49, 59) |
| ≥7 days after Dose 2 | 26 / 21,918 | 2,005 / 119,081 | 93 (90, 95) | 92 (88, 95) |
| No | | | | |
| ≥14 days after Dose 1 | 228 / 29,556 | 3,657 / 138,122 | 71 (67, 75) | 69 (64, 73) |
| ≥7 days after Dose 2 | 31 / 29,359 | 1,812 / 136,277 | 92 (89, 95) | 91 (86, 93) |
| **Epidemic wave** | | | | |
| 14 Dec 2020–7 Feb 2021 | | | | |
| ≥14 days after Dose 1 | Suppressed[c] | 817 / 103,936 | 73 (63, 80) | 62 (48, 73) |
| ≥7 days after Dose 2 | Suppressed[c] | 141 / 103,260 | 96 (73, 100) | 94 (60, 99) |
| 8 Feb 2021–21 Mar 2021 | | | | |
| ≥14 days after Dose 1 | Suppressed[c] | 1,124 / 84,426 | 72 (63, 79) | 61 (46, 71) |
| ≥7 days after Dose 2 | Suppressed[c] | 1,721 / 85,023 | 94 (90, 96) | 92 (86, 95) |
| 22 Mar 2021–19 Apr 2021 | | | | |
| ≥14 days after Dose 1 | 599 / 20,684 | 6,504 / 71,624 | 70 (68, 73) | 59 (55, 62) |
| ≥7 days after Dose 2 | 41 / 20,126 | 1,955 / 67,075 | 93 (91, 95) | 90 (86, 93) |
| **SARS-CoV-2 lineage** | | | | |
| Earlier variant | | | | |
| ≥14 days after Dose 1 | 132 / 27,642 | 8,445 / 259,986 | 86 (83, 88) | 61 (53, 67) |
| ≥7 days after Dose 2 | 13 / 27,523 | 3,817 / 255,358 | 97 (95, 98) | 93 (87, 96) |
| Alpha (B.1.1.7) | | | | |
| ≥14 days after Dose 1 | 300 / 12,582 | 8,445 / 259,986 | 27 (18, 35) | 61 (56, 66) |
| ≥7 days after Dose 2 | 23 / 12,305 | 3,817 / 255,358 | 88 (81, 92) | 90 (85, 94) |
| Beta (B.1.351) or Gamma (P.1) | | | | |
| ≥14 days after Dose 1 | Suppressed[c] | 8,445 / 259,986 | 1 (-35, 27) | 43 (22, 59) |
| ≥7 days after Dose 2 | Suppressed[c] | 3,817 / 255,358 | 85 (52, 95) | 88 (61, 96) |

[a]The denominator represents the total number of test-positive cases or test-negative controls (vaccinated and unvaccinated), whereas the numerator represents only the number of test-positive cases or test-negative controls that are vaccinated.

CONFIDENTIAL

[b]Models were adjusted for age, sex, public health unit region, biweekly period of test, number of SARS-CoV-2 tests in the 3 months prior to 14 December 2020, presence of any comorbidity that increase the risk of severe COVID-19, receipt of influenza vaccination in current or prior influenza season, and neighbourhood-level household income, persons per dwelling, proportion of persons employed as non-health essential workers, and self-identified visible minority quintiles.

[c]Counts of vaccinated test-positive cases for each interval are suppressed because they are either small cell counts (≤5, which cannot be disclosed because of privacy and data obligations) or they could be derived using other estimates in this table.

CONFIDENTIAL

**eTable 7:** Unadjusted and adjusted vaccine effectiveness estimates after Dose 1 (for individuals who received only 1 dose) and Dose 2, by various factors, including vaccine product, patient characteristics, epidemic wave, and SARS-CoV-2 lineage against severe outcomes (hospitalization or death) of symptomatic SARS-CoV-2 infection between 14 December 2020 and 19 April 2021 in Ontario, Canada.

| | Test-positive cases, No. Vaccinated/Total[a] | Test-negative controls, No. Vaccinated/Total[a] | Unadjusted VE% (95% CI) | Adjusted[b] VE% (95% CI) |
|---|---|---|---|---|
| **Vaccine product** | | | | |
| BNT162b2 | | | | |
| ≥14 days after Dose 1 | Suppressed[c] | 7,483 / 259,024 | 26 (2, 44) | 69 (59, 77) |
| After Dose 2[d] | Suppressed[c] | 4,112 / 255,653 | 95 (79, 99) | 96 (82, 99) |
| *By age group* | | | | |
| ≥70 years | | | | |
| ≥14 days after Dose 1 | Suppressed[c] | 1623 / 21904 | 37 (15, 54) | 68 (54, 77) |
| After Dose 2[d] | Suppressed[c] | 458 / 20739 | 95 (66, 99) | 97 (77, 100) |
| 40–69 years | | | | |
| ≥14 days after Dose 1 | Suppressed[c] | 3035 / 109737 | 85 (64, 94) | 84 (60, 93) |
| After Dose 2[d] | Suppressed[c] | 1786 / 108488 | 95 (64, 99) | 93 (48, 99) |
| 16–39 years | | | | |
| ≥14 days after Dose 1 | 0 / 214 | 2825 / 127383 | 100 | 100 |
| After Dose 2[d] | 0 / 214 | 1868 / 126426 | 100 | 100 |
| mRNA-1273 | | | | |
| ≥14 days after Dose 1 | Suppressed[c] | 962 / 252,503 | 21 (-67, 62) | 73 (42, 87) |
| After Dose 2 | Suppressed[c] | 709 / 252,250 | 85 (-9, 98) | 96 (74, 100) |
| *By age group* | | | | |
| ≥70 years | | | | |
| ≥14 days after Dose 1 | Suppressed[c] | 354 / 20635 | 56 (7, 79) | 69 (33, 86) |
| After Dose 2[d] | Suppressed[c] | 404 / 20685 | 95 (61, 99) | 96 (73, 99) |
| 40–69 years | | | | |
| ≥14 days after Dose 1 | Suppressed[c] | 346 / 107048 | 100 | 100 |
| After Dose 2[d] | Suppressed[c] | 194 / 106896 | 100 | 100 |
| 16–39 years | | | | |
| ≥14 days after Dose 1 | 0 / 214 | 262 / 124820 | 100 | 100 |
| After Dose 2[d] | 0 / 214 | 111 / 124669 | 100 | 100 |
| **Age group** | | | | |
| ≥70 years | | | | |
| ≥14 days after Dose 1 | Suppressed[c] | 1,977 / 22,258 | 41 (22, 55) | 67 (54, 76) |
| After Dose 2[d] | Suppressed[c] | 862 / 21,143 | 95 (79, 99) | 97 (86, 99) |

CONFIDENTIAL

| | Test-positive cases, No. Vaccinated/Total[a] | Test-negative controls, No. Vaccinated/Total[a] | Unadjusted VE% (95% CI) | Adjusted[b] VE% (95% CI) |
|---|---|---|---|---|
| 40–69 years | | | | |
| ≥14 days after Dose 1 | Suppressed[c] | 3,381 / 110,083 | 87 (68, 94) | 86 (66, 94) |
| After Dose 2[d] | Suppressed[c] | 1,980 / 108,682 | 95 (68, 99) | 94 (56, 99) |
| 16–39 years | | | | |
| ≥14 days after Dose 1 | 0 / 214 | 3,087 / 127,645 | 100 | 100 |
| After Dose 2[d] | 0 / 214 | 1,979 / 126,537 | 100 | 100 |
| **Sex** | | | | |
| Male | | | | |
| ≥14 days after Dose 1 | Suppressed[c] | 2,317 / 109,592 | 9 (-35, 38) | 72 (57, 81) |
| After Dose 2[d] | Suppressed[c] | 1,071 / 108,346 | 77 (29, 93) | 89 (65, 97) |
| Female | | | | |
| ≥14 days after Dose 1 | 32 / 1,025 | 6,128 / 150,394 | 24 (-8, 47) | 67 (52, 77) |
| After Dose 2[d] | 0 / 993 | 3,750 / 148,016 | 100 | 100 |
| **Presence of any comorbidity** | | | | |
| Yes | | | | |
| ≥14 days after Dose 1 | Suppressed[c] | 4,788 / 121,864 | 26 (3, 43) | 68 (57, 76) |
| After Dose 2[d] | Suppressed[c] | 2,527 / 119,603 | 93 (77, 98) | 96 (86, 99) |
| No | | | | |
| ≥14 days after Dose 1 | Suppressed[c] | 3,657 / 138,122 | 84 (37, 96) | 89 (54, 97) |
| After Dose 2[d] | Suppressed[c] | 2,294 / 136,759 | 100 | 100 |
| **Epidemic wave** | | | | |
| 14 Dec 2020–7 Feb 2021 | | | | |
| ≥14 days after Dose 1 | Suppressed[c] | 817 / 103,936 | 75 (-1, 94) | 71 (-20, 93) |
| After Dose 2[d] | Suppressed[c] | 563 / 103,682 | 100 | 100 |
| 8 Feb 2020–21 Mar 2021 | | | | |
| ≥14 days after Dose 1 | Suppressed[c] | 1,124 / 84,426 | 63 (-15, 88) | 69 (4, 90) |
| After Dose 2[d] | Suppressed[c] | 2,218 / 85,520 | 88 (50, 97) | 90 (59, 98) |
| 22 Mar 2021–19 Apr 2021 | | | | |
| ≥14 days after Dose 1 | Suppressed[c] | 6,504 / 71,624 | 25 (0, 43) | 64 (50, 73) |
| After Dose 2[d] | Suppressed[c] | 2,040 / 67,160 | 95 (68, 99) | 97 (80, 100) |
| **SARS-CoV-2 lineage** | | | | |
| Earlier variant | | | | |
| ≥14 days after Dose 1 | Suppressed[c] | 8,445 / 259,986 | 87 (70, 94) | 76 (46, 90) |
| After Dose 2[d] | Suppressed[c] | 4,821 / 256,362 | 92 (69, 98) | 90 (61, 98) |
| Alpha (B.1.1.7) | | | | |
| ≥14 days after Dose 1 | Suppressed[c] | 8,445 / 259,986 | -77 (-158, -22) | 59 (39, 73) |
| After Dose 2[d] | Suppressed[c] | 4,821 / 256,362 | 89 (24, 99) | 94 (59, 99) |

CONFIDENTIAL

| | Test-positive cases, No. Vaccinated/Total[a] | Test-negative controls, No. Vaccinated/Total[a] | Unadjusted VE% (95% CI) | Adjusted[b] VE% (95% CI) |
|---|---|---|---|---|
| Beta (B.1.351) or Gamma (P.1) | | | | |
| ≥14 days after Dose 1 | 6 / 92 | 8,445 / 259,986 | -108 (-376, 9) | 56 (-9, 82) |
| After Dose 2[d] | 0 / 86 | 4,821 / 256,362 | 100 | 100 |

[a]The denominator represents the total number of test-positive cases or test-negative controls (vaccinated and unvaccinated), whereas the numerator represents only the number of test-positive cases or test-negative controls that are vaccinated.
[b]Models were adjusted for age, sex, public health unit region, biweekly period of test, number of SARS-CoV-2 tests in the 3 months prior to 14 December 2020, presence of any comorbidity that increase the risk of severe COVID-19, receipt of influenza vaccination in current or prior influenza season, and neighbourhood-level household income, persons per dwelling, proportion of persons employed as non-health essential workers, and self-identified visible minority quintiles.
[c]Counts of vaccinated test-positive cases for each interval are suppressed because they are either small cell counts (≤5, which cannot be disclosed because of privacy and data obligations) or they could be derived using other estimates in this table. Overall, 61/2,479 (2.5%) of test-positive cases with a severe outcome were vaccinated.
[d]Due to the small number of severe outcomes after Dose 2, we evaluated VE for the entire period (i.e., ≥0 days) after receipt of the second dose.

CONFIDENTIAL

**eTable 8:** Unadjusted and adjusted vaccine effectiveness estimates against symptomatic SARS-CoV-2 infection and severe outcomes (hospitalization or death), stratified by age group, between 14 December 2020 and 19 April 2021 in Ontario, Canada.

| | Test-positive cases, No. Vaccinated/Total[a] | Test-negative controls, No. Vaccinated/Total[a] | Unadjusted VE% (95% CI) | Adjusted[b] VE% (95% CI) |
|---|---|---|---|---|
| **Symptomatic SARS-CoV-2 infection** | | | | |
| **≥70 years** | | | | |
| **Received one dose, time since Dose 1** | | | | |
| 0–13 days | Suppressed[c] | 1,638 / 21,919 | -52 (-71, -35) | -9 (-26, 6) |
| 14–20 days | Suppressed[c] | 851 / 21,132 | -25 (-49, -5) | 24 (7, 38) |
| 21–27 days | Suppressed[c] | 557 / 20,838 | 9 (-17, 28) | 40 (21, 54) |
| 28–34 days | Suppressed[c] | 322 / 20,603 | 42 (14, 60) | 64 (46, 76) |
| 35–41 days | Suppressed[c] | 177 / 20,458 | 28 (-16, 55) | 64 (41, 78) |
| 42–48 days | Suppressed[c] | 51 / 20,332 | 74 (-8, 94) | 85 (38, 97) |
| ≥49 days | Suppressed[c] | 19 / 20,300 | 65 (-164, 95) | 81 (-48, 98) |
| **Received two doses, time since Dose 2** | | | | |
| ≥7 days | Suppressed[c] | 690 / 20,971 | 93 (86, 97) | 94 (87, 97) |
| **40–69 years** | | | | |
| **Received one dose, time since Dose 1** | | | | |
| 0–13 days | 647 / 24,107 | 2,378 / 109,080 | -24 (-35, -13) | -16 (-27, -5) |
| 14–20 days | 93 / 23,553 | 1,059 / 107,761 | 60 (51, 68) | 54 (42, 63) |
| 21–27 days | 52 / 23,512 | 801 / 107,503 | 71 (61, 78) | 66 (54, 74) |
| 28–34 days | 33 / 23,493 | 685 / 107,387 | 78 (69, 85) | 76 (65, 83) |
| 35–41 days | 31 / 23,491 | 444 / 107,146 | 68 (54, 78) | 71 (58, 80) |
| 42–48 days | 19 / 23,479 | 306 / 107,008 | 72 (55, 82) | 75 (60, 85) |
| ≥49 days | 9 / 23,469 | 86 / 106,788 | 52 (5, 76) | 59 (17, 80) |
| **Received two doses, time since Dose 2** | | | | |
| ≥7 days | 31 / 23,491 | 1,574 / 108,276 | 91 (87, 94) | 89 (85, 93) |
| **16–39 years** | | | | |
| **Received one dose, time since Dose 1** | | | | |
| 0–13 days | Suppressed[c] | 1,940 / 126,498 | 29 (19, 37) | 10 (-4, 21) |
| 14–20 days | Suppressed[c] | 884 / 125,442 | 82 (75, 88) | 78 (68, 85) |

CONFIDENTIAL

| | Test-positive cases, No. Vaccinated/Total[a] | Test-negative controls, No. Vaccinated/Total[a] | Unadjusted VE% (95% CI) | Adjusted[b] VE% (95% CI) |
|---|---|---|---|---|
| 21–27 days | Suppressed[c] | 772 / 125,330 | 79 (70, 85) | 75 (64, 82) |
| 28–34 days | Suppressed[c] | 640 / 125,198 | 63 (50, 73) | 61 (47, 71) |
| 35–41 days | Suppressed[c] | 442 / 125,000 | 75 (62, 84) | 76 (63, 85) |
| 42–48 days | Suppressed[c] | 237 / 124,795 | 55 (30, 72) | 56 (31, 72) |
| ≥49 days | Suppressed[c] | 112 / 124,670 | 60 (20, 80) | 65 (30, 82) |
| **Received two doses, time since Dose 2** | | | | |
| ≥7 days | Suppressed[c] | 1,553 / 126,111 | 94 (90, 96) | 93 (88, 95) |
| **COVID-19 hospitalization or death** | | | | |
| **≥70 years** | | | | |
| **Received one dose, time since Dose 1** | | | | |
| 0–13 days | Suppressed[c] | 1,638 / 21,919 | -8 (-37, 15) | 18 (-7, 37) |
| 14–20 days | Suppressed[c] | 851 / 21,132 | 32 (0, 55) | 58 (35, 72) |
| 21–27 days | Suppressed[c] | 557 / 20,838 | 33 (-10, 59) | 61 (34, 77) |
| 28–34 days | Suppressed[c] | 322 / 20,603 | 45 (-11, 73) | 72 (42, 87) |
| ≥35 days | Suppressed[c] | 247 / 20,528 | 82 (28, 96) | 93 (71, 98) |
| **Received two doses** | | | | |
| After Dose 2[d] | Suppressed[c] | 862 / 21,143 | 95 (79, 99) | 97 (86, 99) |
| **40–69 years** | | | | |
| **Received one dose, time since Dose 1** | | | | |
| 0–13 days | Suppressed[c] | 2,378 / 109,080 | 20 (-23, 48) | 40 (6, 62) |
| 14–20 days | Suppressed[c] | 1,059 / 107,761 | 92 (39, 99) | 92 (41, 99) |
| 21–27 days | Suppressed[c] | 801 / 107,503 | 66 (-5, 89) | 64 (-13, 89) |
| 28–34 days | Suppressed[c] | 685 / 107,387 | 100 | 100 |
| ≥35 days | Suppressed[c] | 836 / 107,538 | 89 (23, 99) | 88 (13, 98) |
| **Received two doses** | | | | |
| After Dose 2[d] | Suppressed[c] | 1,980 / 108,682 | 95 (68, 99) | 94 (56, 99) |
| **16–39 years** | | | | |
| **Received one dose, time since Dose 1** | | | | |
| 0–13 days | Suppressed[c] | 1,940 / 126,498 | Suppressed[†] | -6 (-236, 66) |
| ≥14 days | 0 / 214 | 3,087 / 127,645 | 100 | 100 |
| **Received two doses** | | | | |

CONFIDENTIAL

| | Test-positive cases, No. Vaccinated/Total[a] | Test-negative controls, No. Vaccinated/Total[a] | Unadjusted VE% (95% CI) | Adjusted[b] VE% (95% CI) |
|---|---|---|---|---|
| After Dose 2[d] | 0 / 214 | 1,979 / 126,537 | 100 | 100 |

[a]The denominator represents the total number of test-positive cases or test-negative controls (vaccinated and unvaccinated), whereas the numerator represents only the number of test-positive cases or test-negative controls that are vaccinated.
[b]Models were adjusted for age, sex, public health unit region, biweekly period of test, number of SARS-CoV-2 tests in the 3 months prior to 14 December 2020, presence of any comorbidity that increase the risk of severe COVID-19, receipt of influenza vaccination in current or prior influenza season, and neighbourhood-level household income, persons per dwelling, proportion of persons employed as non-health essential workers, and self-identified visible minority quintiles.
[c]Counts of vaccinated test-positive cases for each interval or unadjusted VE estimates are suppressed because they are either small cell counts (≤5, which cannot be disclosed because of privacy and data obligations) or they could be derived using other estimates in this table.
[d]Due to the small number of severe outcomes after Dose 2, we evaluated VE for the entire period (i.e., ≥0 days) after receipt of the second dose.

CONFIDENTIAL

20 Quinn RR, Laupacis A, Austin PC, *et al.* Using administrative datasets to study outcomes in dialysis

CONFIDENTIAL

patients: a validation study. *Med Care* 2010;**48**:745–50. doi:10.1097/MLR.0b013e3181e419fd
21 Lapointe-Shaw L, Georgie F, Carlone D, *et al.* Identifying cirrhosis, decompensated cirrhosis and hepatocellular carcinoma in health administrative data: A validation study. *PLoS One* 2018;**13**:e0201120. doi:10.1371/journal.pone.0201120
22 Jaakkimainen L, Bronskill SE, Tierney MC, *et al.* Identification of physician-diagnosied Alzheimer's disease and related dementias in population-based administrative data: A validation study using family physicians' electronic medical records. *J Alzheimer's Dis* 2016;**54**:337–49.
23 Tolea MI, Morris JC, Galvin JE. Trajectory of Mobility Decline by Type of Dementia. *Alzheimer Dis Assoc Disord* 2016;**30**:60–6. doi:10.1097/WAD.0000000000000091
24 Korczyn AD. Dementia in the COVID-19 Period. *J Alzheimers Dis* 2020;**75**:1071–2. doi:10.3233/JAD-200609
25 *Dictionary, Census of Population, 2016 - Dissemination area (DA)*. Statistics Canada/Statistique Canada 2016. https://www12.statcan.gc.ca/census-recensement/2016/ref/dict/geo021-eng.cfm (accessed 24 Jul 2021).
26 *Low Income Lines: What they are and how they are created*. Statistics Canada/Statistique Canada 2016. https://www150.statcan.gc.ca/n1/pub/75f0002m/75f0002m2016002-eng.pdf (accessed 24 Jul 2021).
27 *Household size of private household*. Statistics Canada/Statistique Canada https://www23.statcan.gc.ca/imdb/p3Var.pl?Function=DEC&Id=251038 (accessed 24 Jul 2021).
28 *Questionnaire(s) and Reporting guide(s) - Census 2A-L - 2016*. Statistics Canada/Statistique Canada https://www23.statcan.gc.ca/imdb/p3Instr.pl?Function=getInstrumentList&Item_Id=295122&UL=1V& (accessed 24 Jul 2021).

CONFIDENTIAL
